# Shared genetic architecture of stroke and its comorbid conditions reveals latent phenotypes that differ among populations: A genomic structural equation modelling approach

**DOI:** 10.1101/2025.04.11.25325709

**Authors:** Rashmi Sukumaran, Achuthsankar S. Nair, Moinak Banerjee

## Abstract

Stroke affects over 101 million people worldwide and is the second most fatal non-communicable disease with over 6.55 million deaths in 2019. Stroke has a complex etiology, which is further influenced by the genetics of ethnicity and its comorbid conditions. A genomic structure equation model was applied to identify the latent factors for stroke and its comorbid conditions in European and East Asian population. Two latent factors characteristic to the population emerged in each of the populations - one was an immunological factor and other a metabolic factor. The inflammation factor was defined by the shared variants in ischemic stroke and ischemic heart disease in the European population, and by shared variants in ischemic stroke and high systolic blood pressure in the East Asian population. The metabolic factor was defined by the shared variants in high BMI and Type 2 diabetes in both the populations. The significant variants associated with the latent factors were identified using genomic SEM and characterised using FUMA. A total of 99 new loci associated with stroke and its latent factors were identified. Expression quantitative trait loci highlight the differential effect of change in gene expression of the colocalizing variants among the two populations. The drug targets and pathways identified for these latent factors provides direction for drug repurposing and intervention points that can be leveraged to reduce the burden of stroke. Precise understanding of these genomic insights will aid in designing appropriate therapeutic and prevention strategies to reduce the burden of stroke.

## INTRODUCTION

Stroke affects over 101 million people worldwide and is the second most fatal non-communicable disease with over 6.55 million deaths in 2019. Socio-economic differences are often implicated as the root cause for the differences in the burden of stroke seen across regions of the world. Stroke has a complex etiology, which is further influenced by the genetics of ethnicity and the genetics of comorbid conditions of stroke. This was demonstrated in an earlier study that suggested that the genetic distinctions of stroke and its comorbid conditions in the different populations defines the foundation for the differences seen in the burden of stroke [1].

Comorbid conditions have an implication on the outcome of the stroke. Presence of comorbidity can increase the risk for stroke and negatively impact the stroke outcome. Thus, it is pertinent to understand the relationship between stroke and its comorbid conditions. Despite the strong genetic risk that drives both stroke and its comorbid risk conditions, there was minimal sharing of risk loci among the diseases [1]. This led us to the hypothesis that there may be hidden genetic factors influencing stroke and its comorbidities that are unmeasured. Thus, in this work, the aim was to decode the association of latent factors that interact with stroke and its comorbid conditions.

The shared genetic architecture of complex traits and diseases in a population is captured by a key population parameter called genetic correlation. It is the geometric mean of the trait variances based on the genetic variants across the genome. Hence, to decode the association of latent factors that interact with stroke and its comorbid conditions, we employed a flavour of structural equation modelling called Genomic SEM [2]. Structural equation modelling is a statistical method that allows us to define relationships between independent variables and dependent variables. Its aim is to estimate parameters that maximise the likelihood of the observed data, given a model describing the relationship between the variables. The major difference of SEM from other statistical methods, like multiple regression analysis, is that, here the observed data comprises of the observed covariance between the variables and the variance of the variables [3].

## RESULTS

Summary of the cases and the controls used in the study for European and East Asian datasets for stroke and its comorbid conditions is presented in (Supplementary table S1).

### Genetic correlation and heritability of stroke and its comorbid conditions varies among populations

LD score regression (LDSC) method implemented in Genomic SEM software was used to calculate the genetic correlation between stroke and its comorbid conditions using nearly eight million SNPs each from the European as well as East Asian populations, and is shown in Figure 1. In the European population, the highest genetic correlation is observed between ischemic stroke and ischemic heart disease, followed by ischemic stroke with type 2 diabetes and high systolic BP. However, in the East Asian population, the highest genetic correlation is seen between type1 and type 2 diabetes, followed closely by type 1 diabetes and chronic kidney disease. Ischemic stroke has the highest correlation with high systolic BP in the East Asian population. In both the populations, high LDL shows negative or near zero genetic correlation with all other diseases.

**Figure 1.**
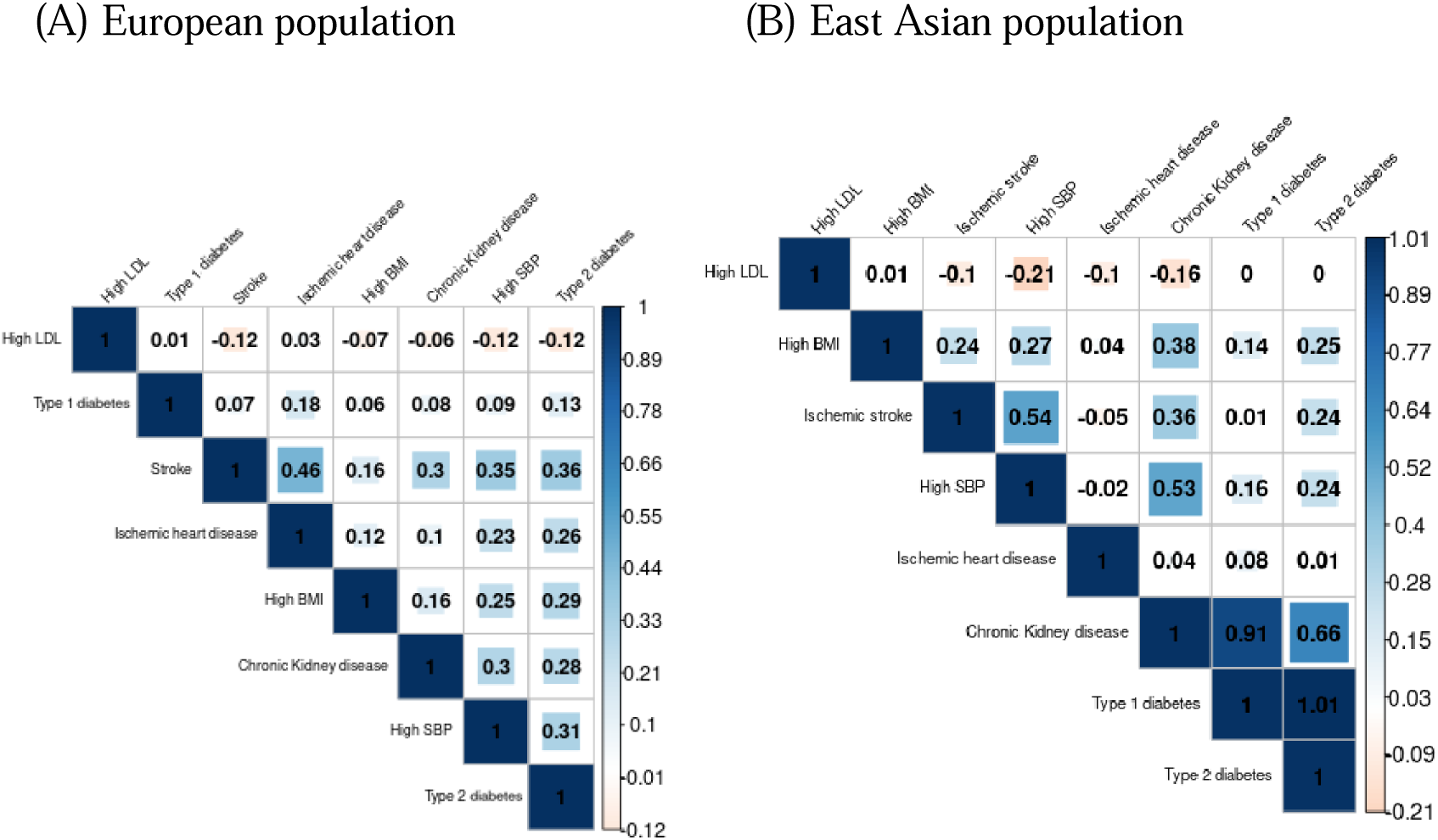
LDSC genetic correlation of stroke and its comorbid conditions in the (A) European and (B) East Asian population. The genetic correlation among stroke and its comorbid conditions calculated using variants in GWAS summary statistics by the LD score regression method implemented in Genomic SEM.

LDSC also calculates the heritability of each individual trait (Supplementary table S2). It is evident that the heritability varies greatly between the populations. Type 1 diabetes that has the highest heritability in the European population, while it has one of the lowest heritability in East Asia. In fact, all the metabolic diseases, except chronic kidney diseases, have heritability higher than stroke, ischemic heart disease and high systolic BP in the European population. On the other hand, the pattern reverses in the East Asian population, where stroke, ischemic heart disease and systolic BP have higher heritability than high LDL cholesterol, type 1 diabetes and chronic kidney disease.

### Structural equation models of stroke and its comorbid conditions varies among populations

Mathematical models that describe the genetic relationship between stroke and its comorbid conditions were defined based on the LDSC genetic correlation, estimated from GWAS summary statistics using genomic SEM. The exploratory factor analysis (EFA) investigated how many latent factors could be present for the traits under investigation. In both the European and East Asian populations, two-factor models were observed to be optimal to describe the relationships (Supplementary figure S1). Confirmatory factor analysis (CFA) was done with one-, two- and three-factor models for completeness for both the populations. Multiple model fit indices showed reliably that the two-factor model suited our data in both the populations. Sample models tested for European and East Asian populations are shown in Supplementary figures S2A and S2B, respectively. Only models with factors loadings of 0.3 or greater on all variables were considered [4].

For the European populations, two models showed excellent model fit. Both models had low χ^2^ goodness-of-fit index equal to 0.015, one degree of freedom, with non-significant p-value of 0.9 (>0.05). Other fit indices included a CFI of one (>0.95) and an SRMR of 0.0017 (<0.05). Both these models were considered as the final proposed genetic models for stroke and its comorbid conditions in the European population. Both the models had two latent factors, one latent factor (F1) defined by ischemic stroke and ischemic heart disease, and the other factor (F2) defined by high BMI and type 2 diabetes. Both factors in the models had path coefficients greater than 0.3 to all the diseases, and all paths were statistically significant (Table 1, Supplementary table S3). The only difference is that in the second model, the second factor (F2) is also defined by the first factor (F1) with a path coefficient of 0.56. The two models were named EUR Stroke Model A (Figure 2) and EUR Stroke Model B (Supplementary figure S3), respectively. The EUR Stroke model A has been elaborated further in all analysis.

**Figure 2.**
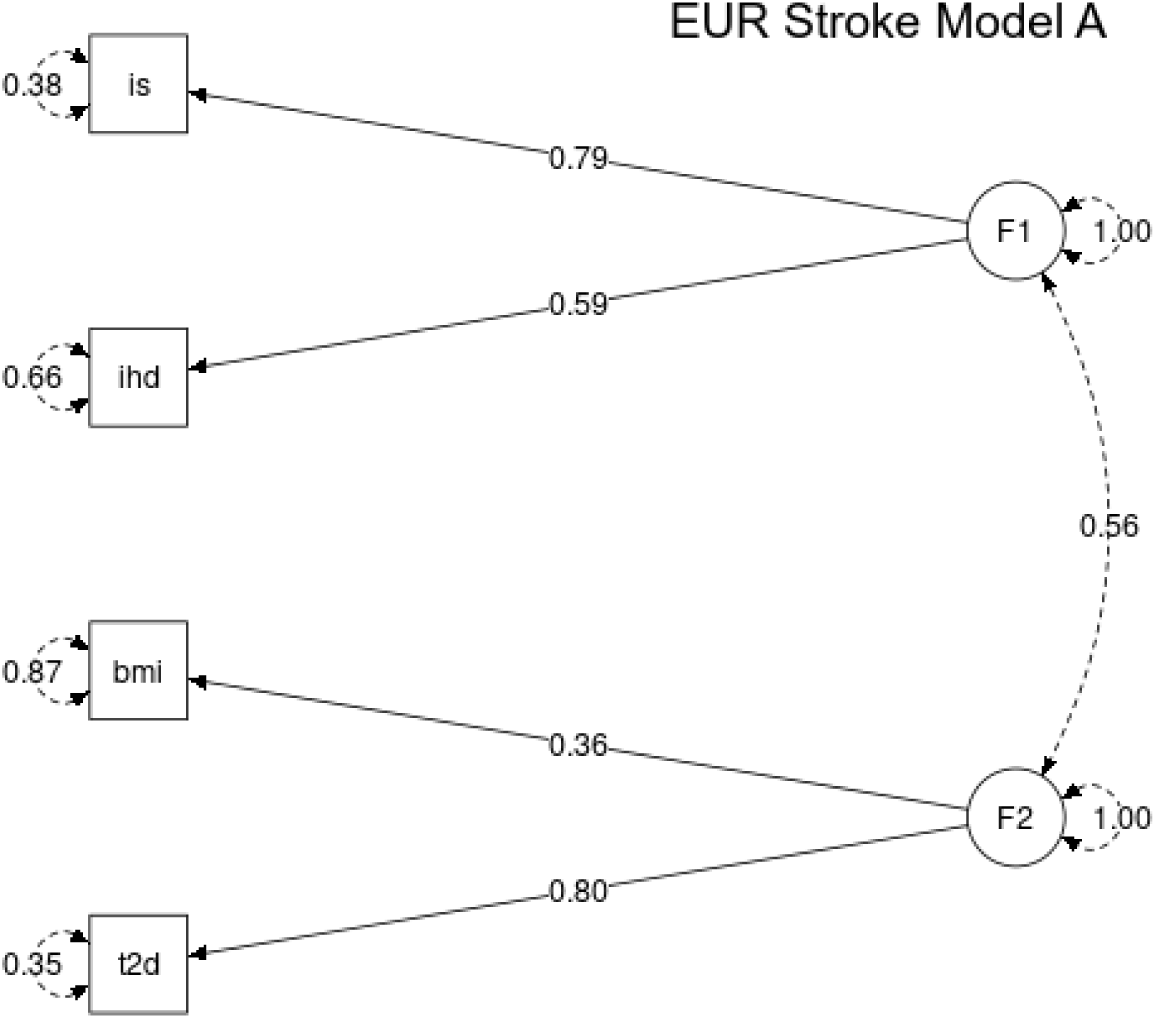
Genetic models proposed for stroke and its comorbid conditions in the European population. The proposed genetic models for stroke and its comorbid conditions have two latent factors. Ischemic stroke (is) and ischemic heart disease (ihd) loads onto factor one, F1, and high body mass index (bmi) and type 2 diabetes (t2d) loads onto second factor F2. Both factors, F1 and F2, have path coefficients to each variable greater than 0.3. In model A, the two factors have a correlation of 0.56.

**Table 1.**
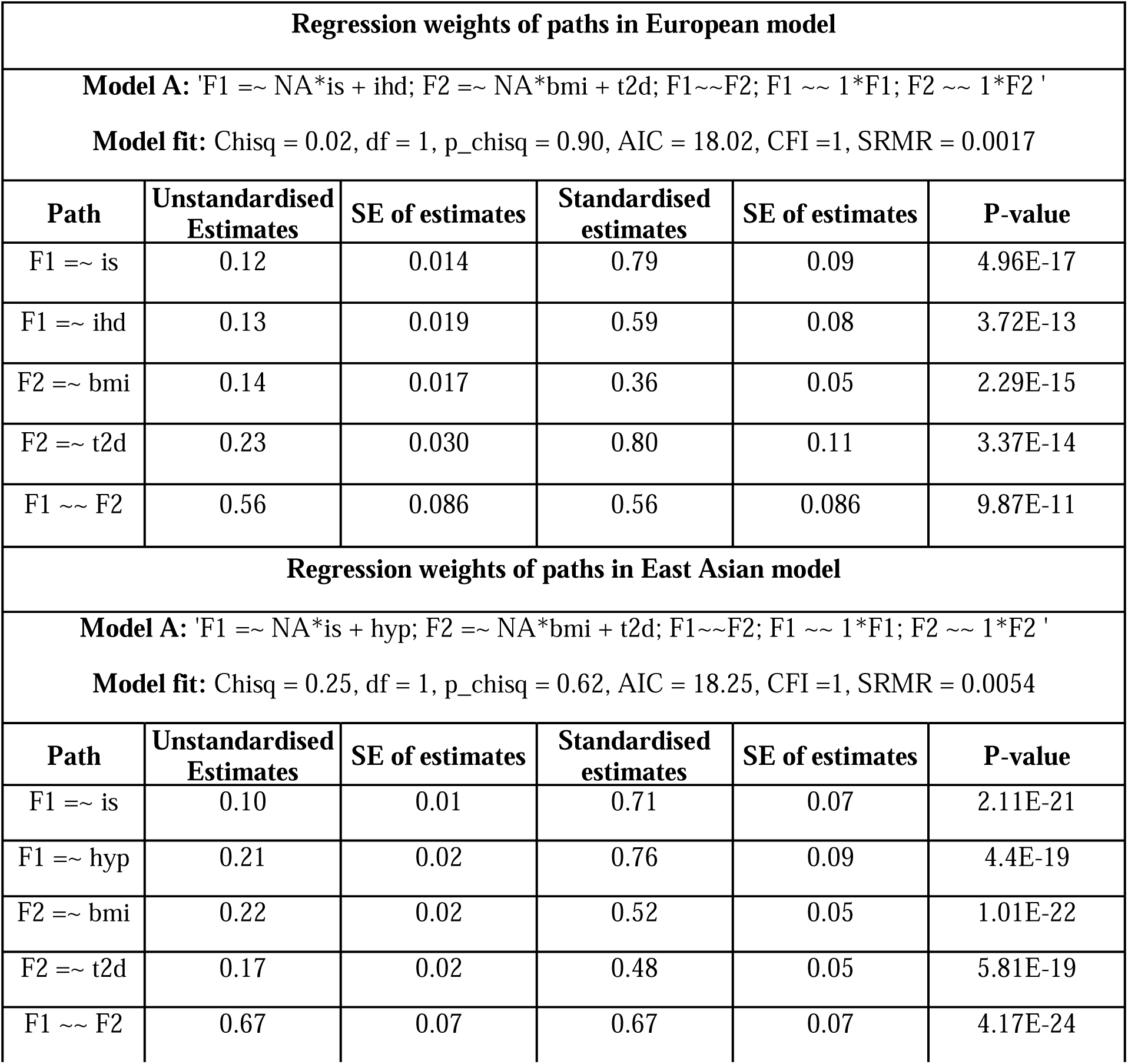
Summary of regression weights of factor loadings in the genetic models in European and East Asian populations. The table shows the paths, their unstandardised and standardised estimates along with p-value. Is - ischemic stroke, ihd - ischemic heart disease, hyp - high systolic BP, bmi - high BMI, t2d - type 2 diabetes.

For the East Asian populations, multiple models showed excellent fit to the data when considering the model fit indices (Supplementary table S3). Four models with one latent factor, five models with two latent factors and one model with three latent factors were observed to have excellent model fit indices. However, when factor loadings on the variables are considered, all models had very poor loading (−0.01) for ischemic heart disease (ihd) as can be seen in Supplementary figure S2B, and the path was not significant (p-value > 0.05). Hence, all the models containing ischemic heart disease were rejected.

The final proposed genetic model for the East Asian population were two models. Both had low χ^2^ goodness-of-fit index equal to 0.25, one degree of freedom, with non-significant p-value of 0.6 (>0.05). Other fit indices included a comparative fit index of one (>0.95) and an SRMR of 0.0054 (<0.05). Both the models had two latent factors, one latent factor (F1) defined by ischemic stroke and high systolic blood pressure, and the other factor (F2) defined by high BMI and type 2 diabetes. Both factors in the models have path coefficients of greater than 0.3 to all the diseases, and all paths are statistically significant (Table 1). The only difference is that in the second model, the first factor (F1) also loads onto the factor two (F2) with a path coefficient of 0.67. The two models were named EAS Stroke Model A (Figure 3) and EAS Stroke Model B (Supplementary figure S4), respectively. The EAS Stroke model A has been elaborated further in all analysis.

**Figure 3.**
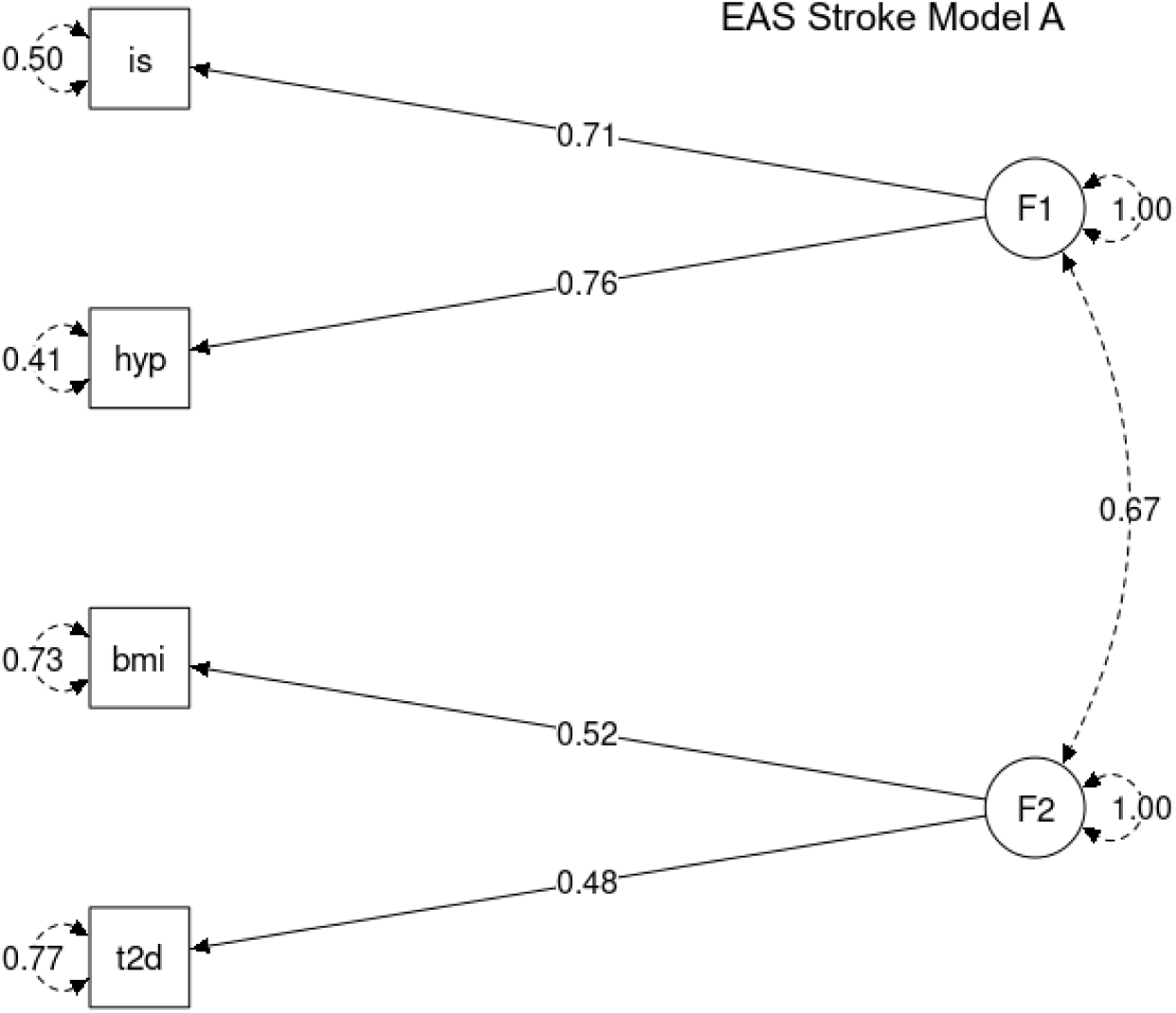
Genetic models proposed for stroke and its comorbid conditions in the East Asian population. The proposed genetic models for stroke and its comorbid conditions have two latent factors. One loads on ischemic stroke (is) and high systolic BP (hyp), and the other loads on high body mass index (bmi) and type 2 diabetes (t2d). Both factors, F1 and F2, have path coefficients to each variable greater than 0.3 in all models. In model A, the two factors have a correlation of 0.67. In model B, factor 2 also loads on factor 1 with a path coefficient of 0.67. The residuals of the variables are shown in circular self-paths.

In both the populations, the proposed genetic models are highly similar. However, a major difference being ischemic stroke and ischemic heart disease loading onto factor one (F1) in the European population, which is replaced by ischemic stroke and high systolic BP loading onto factor one (F1) in the East Asian population. This was as expected from literature, high systolic BP was not expected in the final model in the European population, while it was expected for the East Asian [1]. The factors identified in all models are genetically defined factors that could represent some trait/condition that is unmeasured. These factors summarise the genetic variance shared between the corresponding diseases they represent.

### Genome wide association analysis of the identified latent factors

For each model, genomic SEM was used to do a multivariate genome wide association (GWA) analysis on the estimated latent factors (F1 and F2) to identify SNPs associated with them. Genomic SEM was run on each SNP to estimate its effect on the defined model. SNPs were taken from the individual summary statistics of each trait - 12,064,706 SNPs from the European population and 28,219,311 SNPs from the East Asian population, and were tested on each of the factors in all the models, and summary statistics for each of the factors in all the models were generated.

The summary statistics were analysed with FUMA to identify SNPs significantly associated with each factor. Significant SNPs with p-value ≤ 5×10^-8^ that were independent (LD measure r^2^ < 0.1) were identified as lead SNPs. SNPs in linkage disequilibrium (LD measure r2 ≥ 0.6) and present in the 1000 Genomes reference panel for the concerned population (European or East Asian) were identified as candidate SNPs.

In European stroke Model A, out of a total of 12,064,706 SNPs for the four diseases, 874,673 were found to be associated with first factor F1 (ischemic stroke and ischemic heart disease) and 874,673 were found to be associated with factor two F2 (high BMI and type 2 diabetes). Out of these associated SNPs, a total of 11 genome-wide significant (P< 5e-08) and independent lead SNPs (LD measure r2 < 0.1) were identified for factor F1 (Table 2). Out of these 11 SNPs, five are novel and have not been reported in GWAS Catalog for either ischemic stroke or ischemic heart disease. Additionally, a total of 1083 SNPs were statistically significant and in linkage disequilibrium with the 11 lead SNPs, and were termed as candidate SNPs (Supplementary Figure S5, top panel). In the same model, a total of 14 genome-wide significant and independent SNPs were identified for factor F2, onto which high BMI and type 2 diabetes loads (Table 2). Out of these 14 lead SNPs, three are novel and have not been reported in GWAS Catalog for either high BMI or type 2 diabetes. A total of 1531 SNPs in linkage disequilibrium with these 14 SNPs identified were also statistically significant and termed as candidate SNPs (Supplementary Figure S5, bottom panel).

**Table 2.** Significant lead SNPs associated with latent factors of stroke and its comorbid conditions in the European population. The table shows the statistically significant (P<5e-08) and independent (LD measure r^2^ < 0.1) SNPs associated with each factor. Annotated gene is the gene which has at least one functional annotation with respect to the SNP. Nearest gene is the gene physically closest to the SNP. GWAS reported trait shows if the SNP has been reported for stroke or its comorbid conditions.

| **Factor 1: F1 =~ NA\*is + ihd** |  |  |  |  |  |
| --- | --- | --- | --- | --- | --- |
| No | SNP | Location | Mapped gene | Novel | GWAS Reported trait |
| 1 | rs1448817 | 4:111641053 | PITX2 | Yes | - |
| 2 | rs2066861 | 4:155527436 | FGG | Yes | - |
| 3 | rs4977574 | 9:22098574 | CDKN2B | No | IHD |
| 4 | rs495828 | 9:136154867 | ABO | No | IHD |
| 5 | rs10774625 | 12:111910219 | SH2B3 | No | Large artery stroke, IHD |
| 6 | rs4932370 | 15:91404705 | FURIN | No | Stroke |
| 7 | rs11656775 | 17:17654319 | RAI1 | Yes | - |
| 8 | rs17608766 | 17:45013271 | GOSR2 | No | IHD |
| 9 | rs1560711 | 19:10742287 | ILF3 | Yes | - |
| 10 | rs1122608 | 19:11163601 | LDLR | No | Large artery stroke, IHD |
| 11 | rs17092215 | 20:33595913 | GSS | Yes | - |
| **Factor 2: F2 =~ NA\*bmi + t2d** |  |  |  |  |  |
| No | SNP | Location | Mapped gene | Novel | GWAS Reported trait |
| 1 | rs2012697 | 1:72819612 | NEGR1 | Yes | - |
| 2 | rs543874 | 1:177889480 | SEC16B | No | BMI |
| 3 | rs4854344 | 2:638144 | TMEM18 | No | BMI |
| 4 | rs7647305 | 3:185834290 | ETV5 | No | BMI |
| 5 | rs13130484 | 4:45175691 | GNPDA2 | No | BMI, Type 2 diabetes |
| 6 | rs987237 | 6:50803050 | TFAP2B | No | BMI |
| 7 | rs4074134 | 11:27647285 | BDNF | Yes | - |
| 8 | rs7124681 | 11:47529947 | NDUFS3 | No | BMI, Type 2 diabetes |
| 9 | rs7138803 | 12:50247468 | BCDIN3D | No | BMI |
| 10 | rs7141420 | 14:79899454 | DIO2 | No | BMI |
| 11 | rs11639988 | 16:19944363 | GPRC5B | No | BMI |
| 12 | rs17817449 | 16:53813367 | RPGRIP1L | No | BMI |
| 13 | rs571312 | 18:57839769 | MC4R | No | BMI |
| 14 | rs9675886 | 18:57969322 | MC4R | Yes | - |

In East Asian stroke Model A, out of a total of 28,219,311 SNPs for the four diseases, 5,977,884 were found to be associated with first factor F1 (ischemic stroke and high systolic BP) and 5,977.884 were found to be associated with factor two F2 (high BMI and type 2 diabetes). For factor one, F1, a total of 19 genome-wide significant and independent lead SNPs were identified (Table 3). Out of these 19 SNPs, three are novel and have not been reported in GWAS Catalog, for either ischemic stroke or high systolic BP. A total of 1583 SNPs were in linkage disequilibrium with these 19 SNPs were termed as candidate SNPs (Supplementary Figure S6, top panel). In the same model, factor F2, onto which high BMI and type 2 diabetes, a total of 150 genome-wide significant and independent SNPs were identified (Table 3). Out of these 150 SNPs, 88 are novel and have not been reported in GWAS Catalog for either high BMI or type 2 diabetes. A total of 14,296 SNPs were in linkage disequilibrium with these 150 SNPs and termed as candidate SNPs (Supplementary figure S6, bottom panel). Though the models are similar in both the populations, interestingly, there are no overlapping lead SNPs for any two factors.

**Table 3.**
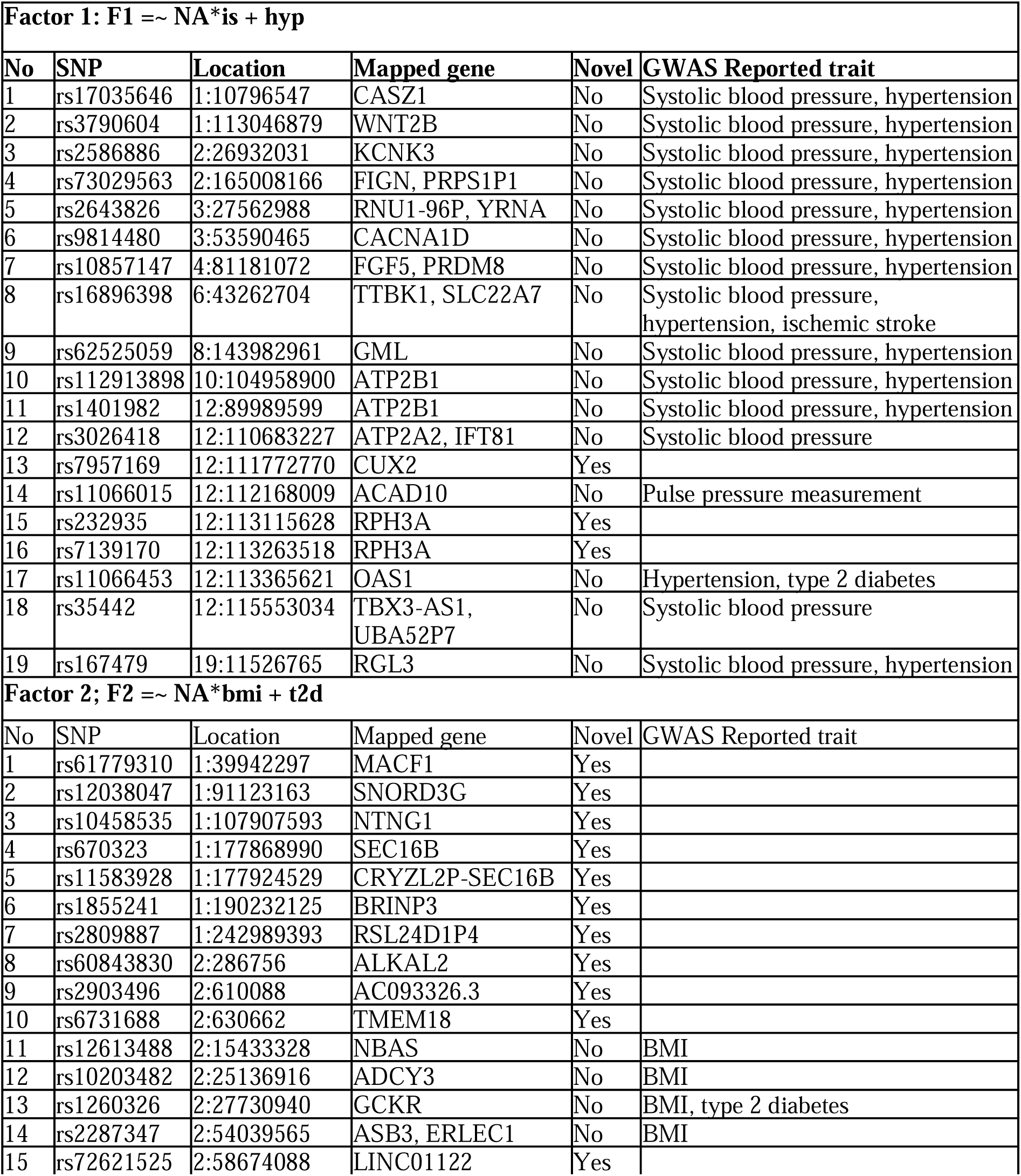
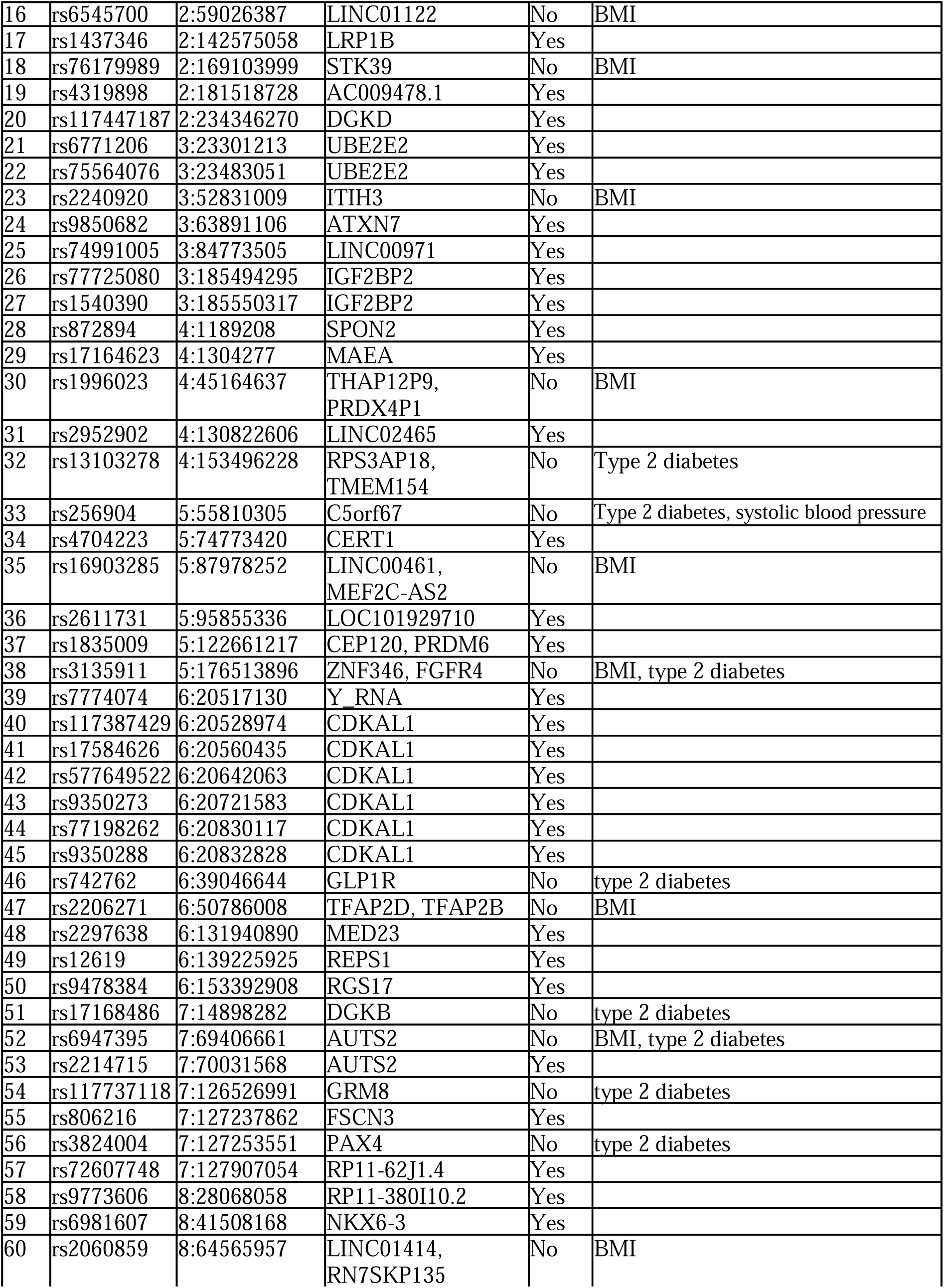
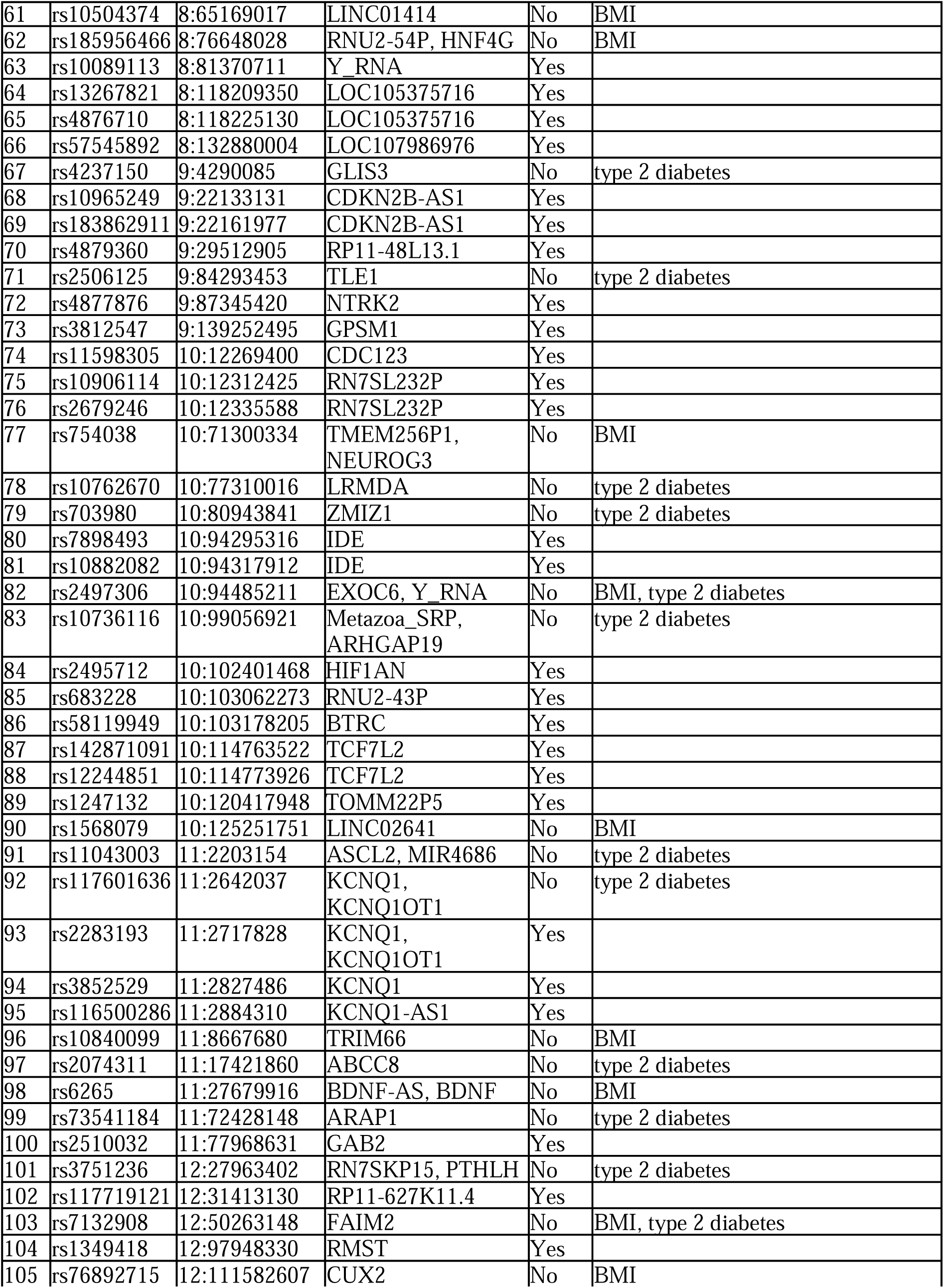
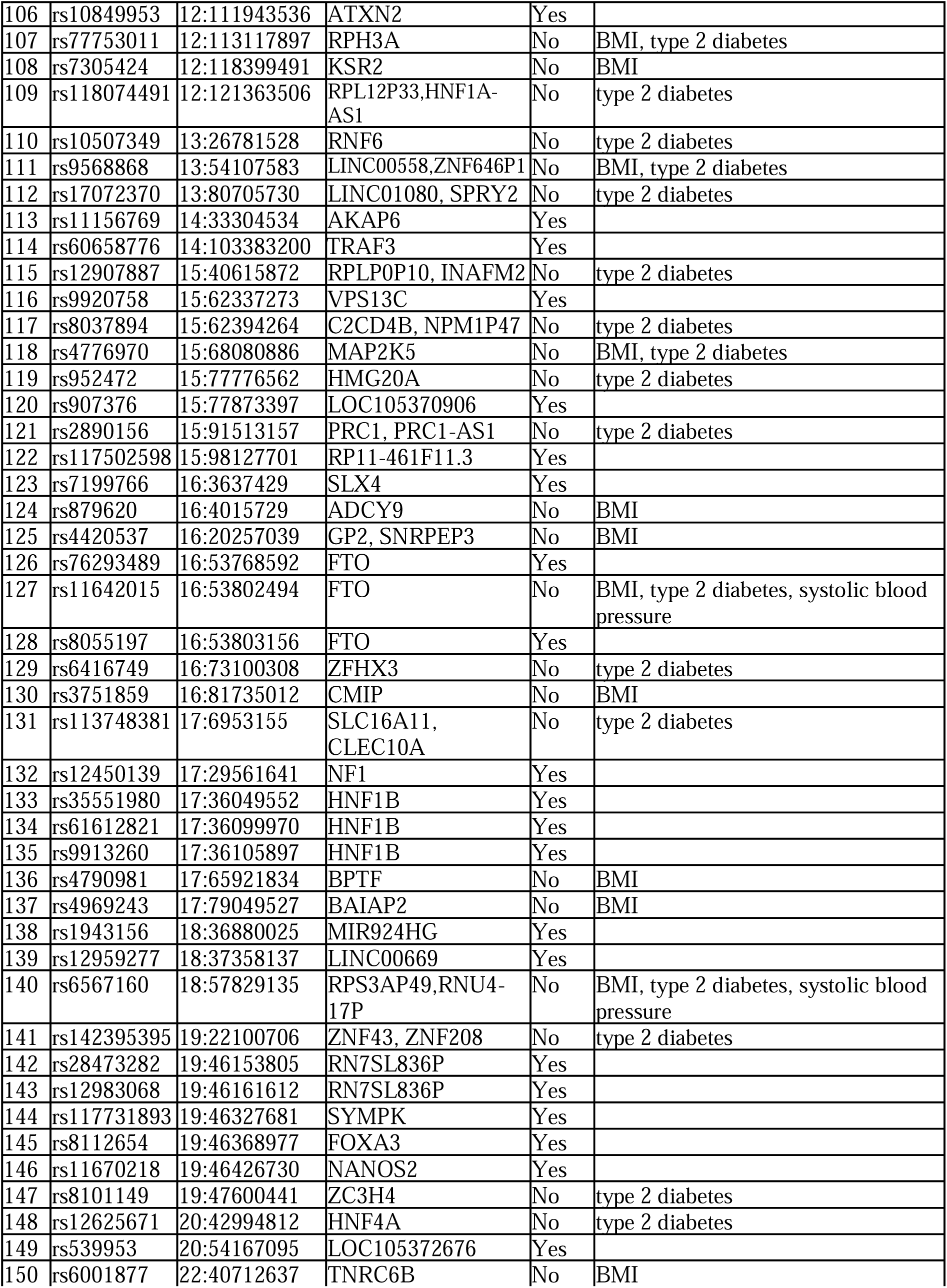
Significant lead SNPs associated with latent factors of stroke and its comorbid conditions in the East Asian population. The table shows the statistically significant (P<5e-08) and independent (LD measure r^2^ < 0.1) SNPs associated with each factor. Annotated gene is the gene which has at least one functional annotation with respect to the SNP. Nearest gene is the gene physically closest to the SNP. GWAS reported trait shows if the SNP has been reported for stroke or its comorbid conditions.

Q_SNP_ heterogeneity tests were done to validate that the lead SNPs act through the model specified and not directly via the individual disease/ traits. For both the European and East Asian model, all lead SNPs for both factors were non-significant (Supplementary Figure S7), indicating that all the SNPs estimated in our models were indeed acting through the model specified.

### Functional Annotation of significant SNPs associated with a factor

The statistically significant SNPs for each factor was annotated using SNP2GENE function in FUMA. FUMA mapped the candidate SNPs to a gene based on position in the human genome assembly GRCh37, as well as with the expression level of the gene based on expression quantitative trait loci (eQTLs).

In European stroke model, based on position (nearest gene less than 10Kb), the 1083 candidate SNPs associated with the first factor F1 (ischemic stroke and ischemic heart disease), had a total of 800 gene mappings (59 unique genes), and the 1531 candidate SNPs in the second factor F2 (high BMI and type 2 diabetes) mapped to a total of 573 genes (36 unique genes). In both the factors, the major proportion of SNPs were in the introns of either protein coding genes or non-coding RNA genes, and in the intergenic regions, the proportions of which are enriched compared to reference proportions, based on ANNOVAR annotations as shown in figure 4.

**Figure 4.**
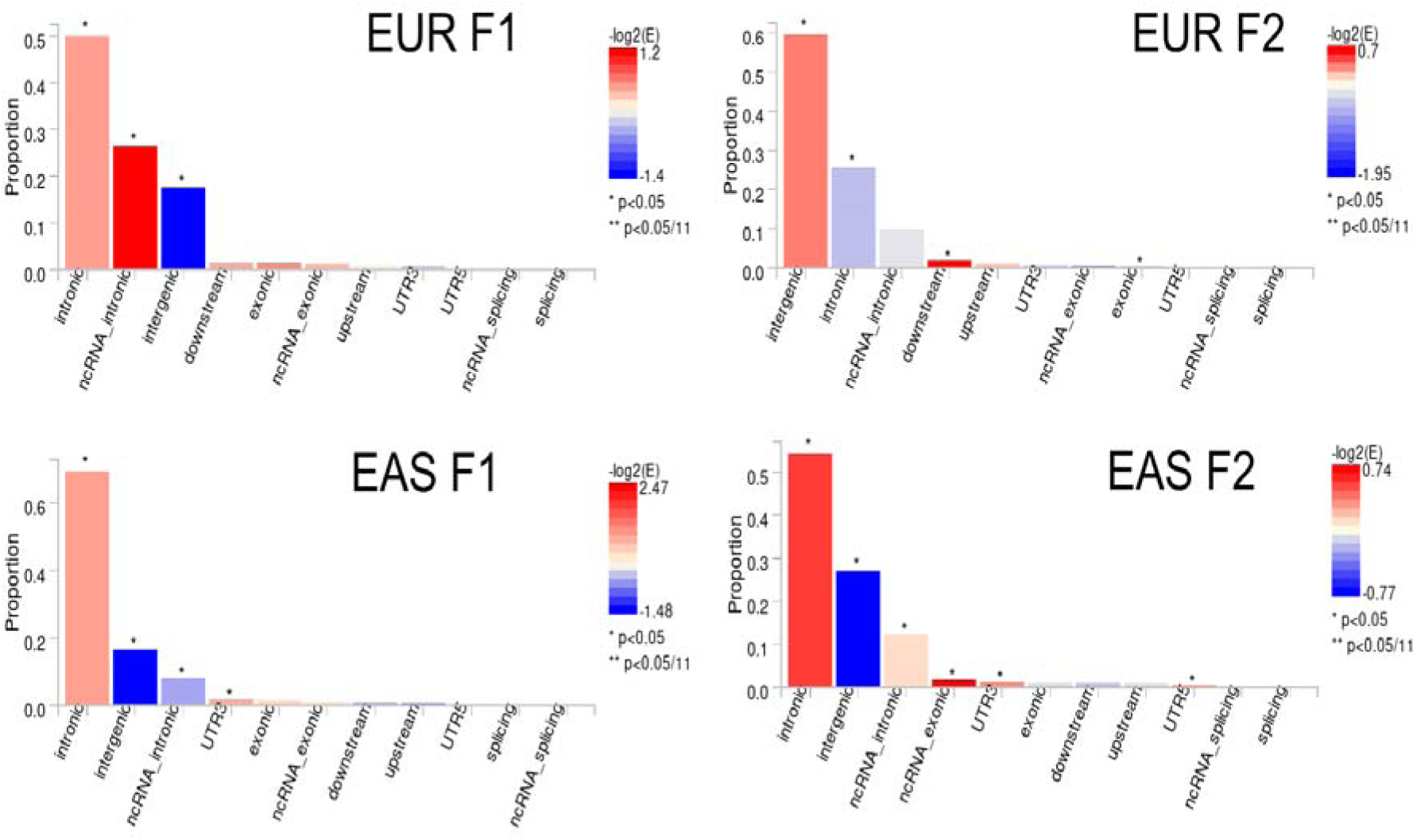
The gene location of the SNPs associated with the latent factors. The histogram displays the proportion of all SNPs in LD with significant SNPs which have corresponding functional annotation from ANNOVAR. Bars are coloured by log2(enrichment) relative to all SNPs selected in the reference panel. EUR F1 and EUR F2 are factors in European population and EAS F1 and EAS F2 are factors in East Asian population. The distribution of exonic, downstream, and UTR annotations are shown separately in Supplementary figure 8.

In East Asian stroke model, based on position, the 1583 candidate SNPs associated with the first factor F1 (ischemic stroke and high systolic BP), mapped to a total of 1317 genes (78 unique genes), and the 14,296 candidate SNPs in the second factor F2 (high BMI and type 2 diabetes) mapped to a total of 10162 genes (294 unique genes). In both the factors, the major proportion of SNPs are in the introns of protein coding genes or of non-coding RNA genes and in the intergenic regions, the proportions of which are enriched compared to reference proportions, based on ANNOVAR annotations as shown in figure 4.

For the eQTL mapping, the candidate SNPs were mapped to loci affecting gene expression in the brain and heart tissues in GTEx v8. In European stroke model, the 1083 candidate SNPs associated with the first factor F1 (ischemic stroke and ischemic heart disease), mapped to a total of 678 genes, whereas the 1531 candidate SNPs in the second factor F2 (high BMI and type 2 diabetes) mapped to a total of 301 genes. In East Asian stroke model, the 1583 candidate SNPs associated with the first factor F1 (ischemic stroke and high systolic BP), mapped to a total of 492 genes, and the 14,296 candidate SNPs in the second factor F2 (high BMI and type 2 diabetes) mapped to a total of 5104 genes.

### Overlapping loci among European and East Asian population shows contrasting expression

For the latent factor F1, there is only one region that overlaps between the two populations. It is present in chromosome 12 and has partial overlap between Europe (111,359,712-112,985,328) and East Asia (110,297,784-113,944,048), as shown in figure 5A and B, respectively. This region comprises 34 significant SNPs associated with F1 in East Asia and 7 significant SNPs associated with F1 in Europe. It also shows significant eQTL expression in different brain and heart tissues for 10 genes associated with F1 in East Asia - *NAA25, ALDH2, FAM216A, RPH3A, RITA1, MAPKAPK5, ACAD10, TMEM116, HECTD4,* and *RAD9B*, and for seven genes associated with F1 in Europe - *ALDH2, NAA25, MAPKAPK5, HECTD4, TMEM116, MYL2,* and *ACAD10*.

**Figure 5A.**
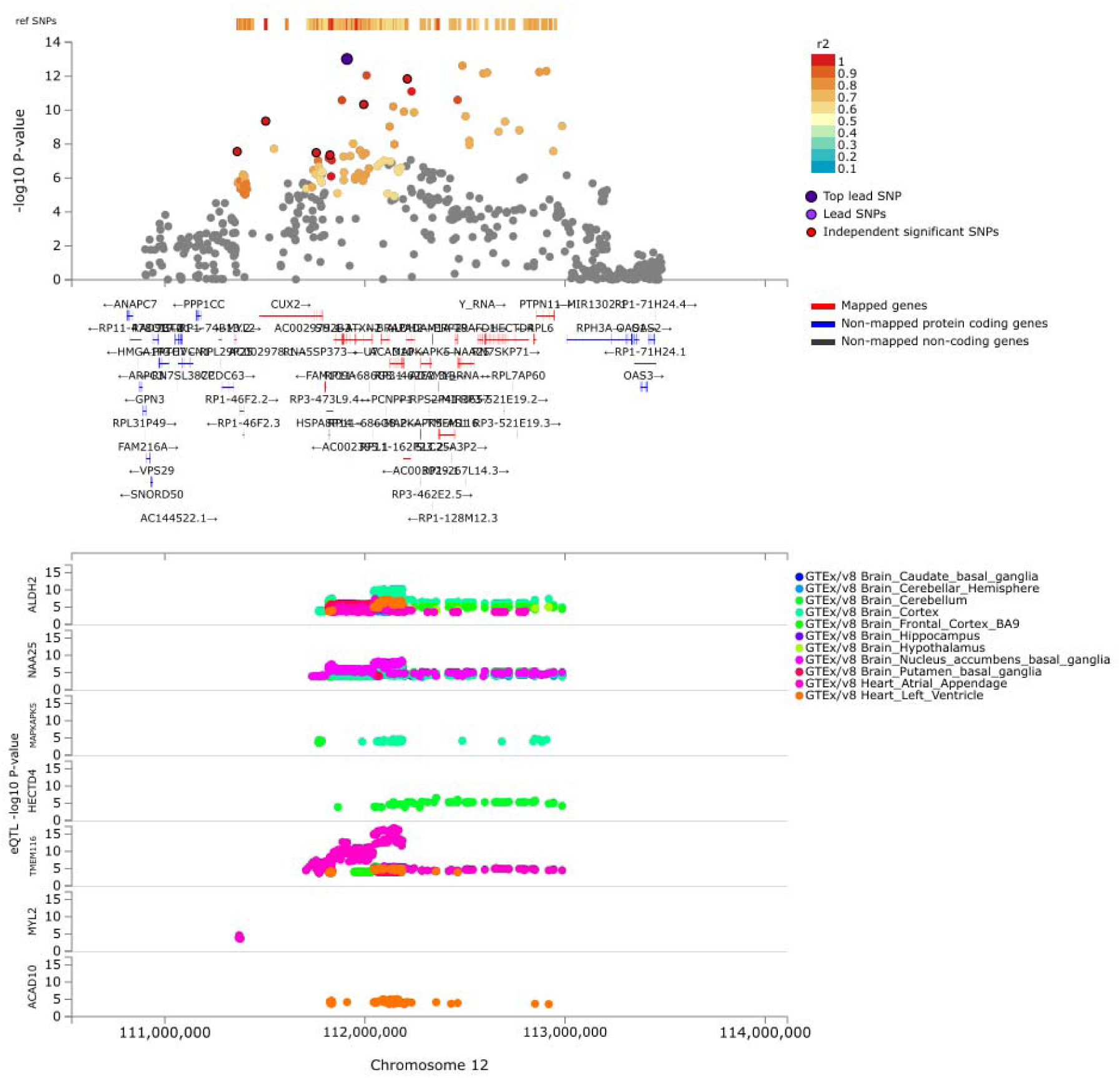
Overlapping genomic risk locus on chromosome 12 associated with latent factor F1 in Europe and East Asia. This is the only genomic risk locus that overlaps between Europe and East Asia for latent factor F1. It comprises 7 significant SNPs from F1 mapping to multiple genes in Europe. It also shows eQTL expression of 7 genes of F1 in different brain and heart tissues.

**Figure 5B.**
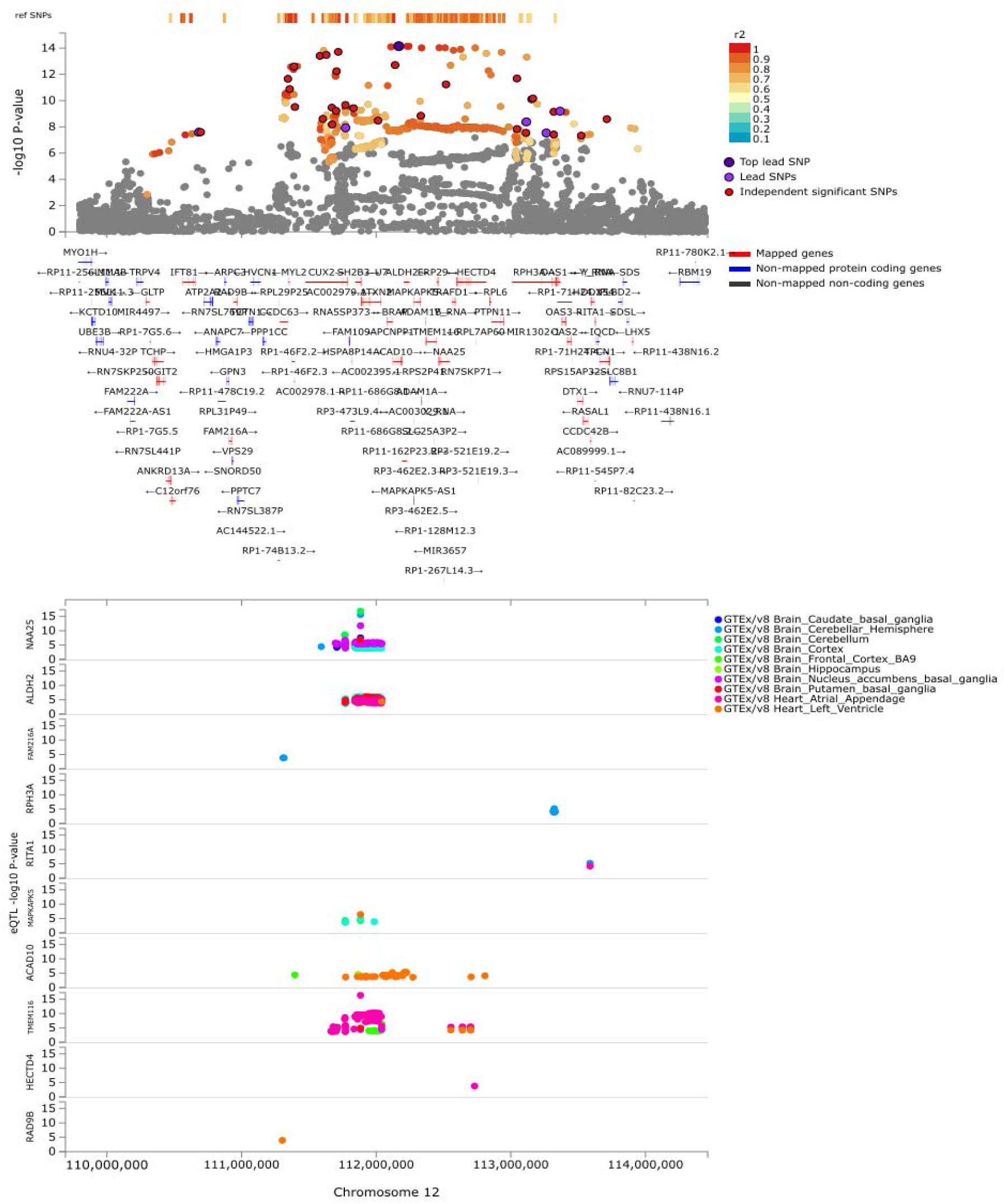
Overlapping genomic risk locus on chromosome 12 associated with latent factor F1 in Europe and East Asia. This is the only genomic risk locus that overlaps between Europe and East Asia for latent factor F1. It comprises 34 significant SNPs from F1 mapping to multiple genes in East Asia. It also shows eQTL expression of 10 genes of F1 in different brain and heart tissues.

Interestingly, though the genes are the same in the two populations, it is evident from eQTL expression in figure 5A and B that the SNPs associated with the factors and their expression in the brain and heart tissue varies contrastingly. This is the direct evidence for the differences in stroke risk among populations going down to the gene-variant level. Similarly, there are seven genomic risk loci overlapping between latent factors F2 in European and East Asian models (Supplementary table S4).

The alignment of the GWAS alleles with the eQTL alleles of the genes in the overlapping loci region is shown in Table 4. For genes *NAA25, ALDH2, MAPKAPK5 and TMEM116*, the alleles in the region have opposite effects on the gene expression in European and East Asian population. The alleles in the European population show decrease in expression of *NAA25* in brain tissues, while the alleles in the East Asian population show increase in expression. The alleles of *ALDH2* show decrease in expression in brain and heart tissues in East Asian population, while the alleles in the European population show increase in expression in both tissues. For the gene *MAPKAPK5*, while the alleles in the East Asian population show decrease in gene expression in heart tissues, while in brain tissue some alleles show decrease in gene expression while some show increase in gene expression. However, in the European population, all alleles of *MAPKAPK5* show increase in gene expression in brain tissue. The *TMEM16* gene also shows differential expression in the populations, while alleles in the European population show decrease in gene expression in both brain and heart, the alleles in the East Asian population show increase in expression in the heart and brain.

**Table 4.** Alignment of GWAS alleles with eQTL alleles in the overlapping genomic risk locus. The table shows the effect of the GWAS risk allele on the expression of genes in brain and heart tissues in the overlapping genomic risk locus in the two populations. Differential expression change in the populations is highlighted in bold.

| Gene | European |  | East Asian |  |
| --- | --- | --- | --- | --- |
|  | # SNPs with eQTL | Change in gene expression | # SNPs with eQTL | Change in gene expression |
| *NAA25* | 191 | Decrease (Brain) | 223 | Increase (Brain,<br>13 loci have decrease) |
| *ALDH2* | 253 | Increase (Heart)<br>Increase (Brain) | 328 | Decrease (Heart)<br>Decrease (Brain,<br>11 loci have increase) |
| *FAM216A* | - | - | 2 | Increase (Brain) |
| *RPH3A* | - | - | 19 | Increase (Brain) |
| *RITA1* | - | - | 2 | Increase (Heart)<br>Increase (Brain) |
| *MAPKAPK5* | 13 | Increase (Brain) | 7 | Decrease (Heart)<br>Increase/Decrease<br>(Brain 3/3 loci) |
| *ACAD10* | 18 | Decrease (Heart) | 40 | Decrease (Heart)<br>Increase (Brain) |
| *TMEM116* | 95 | Decrease (Heart)<br>Decrease (Brain) | 116 | Increase (Heart)<br>Increase (Brain,<br>24 loci have decrease) |
| *HECTD4* | 16 | Increase (Brain) | - | - |
| *RAD9B* | - | - | 1 | Decrease (Heart) |
| *MYL2* | 1 | Decrease (Heart) | - | - |

### Pathway analysis and drug target analysis

In complex traits, genes seldom act in isolation. Hence, pathway analysis was done to elucidate the pathways through which our candidate genes act. Unique genes associated with each factor in both the European and East Asian models were subjected to pathway analysis via ShinyGO server. The pathways with an FDR less than 0.05 were considered significant. Significant pathways associated with each factor, their FDR and fold enrichment compared to background genes are shown in figure 6.

**Figure 6.**
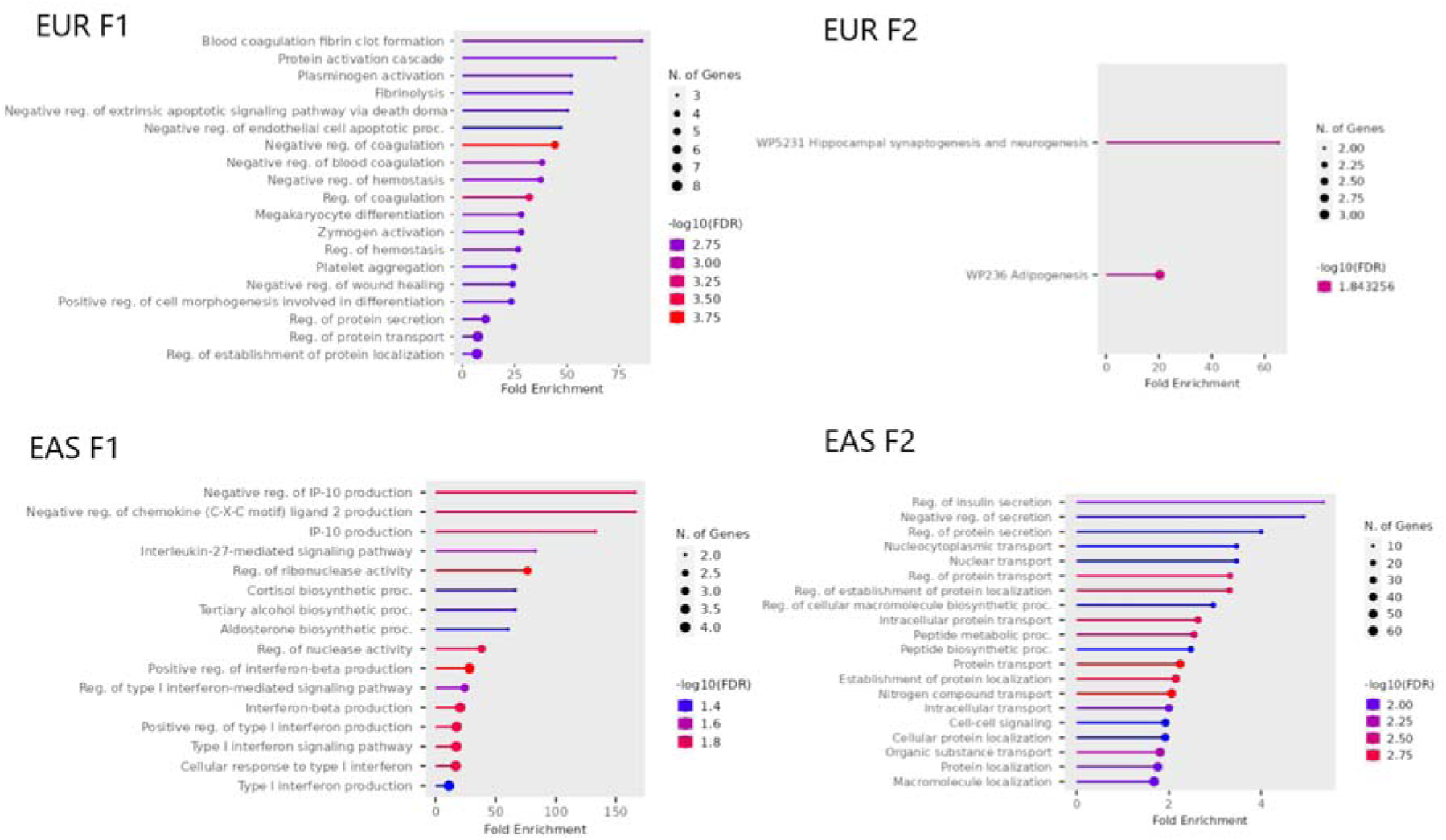
Pathways associated with latent factors in Europe and East Asia. Significantly enriched pathways (FDR < 0.05) associated with each latent factor in the genetic models of Europe and East Asia are shown. The pathways are from databases like KEGG, Reactome, PANTHER, Wiki and GO Biological processes.

In the European genetic model of stroke, the genes associated with latent factor one (F1) are primarily enriched in blood coagulation cascade, protein activation, plasminogen activation etc. with a fold enrichment greater than 30. In the East Asian genetic model of stroke, the genes associated with latent factor one (F1) are primarily enriched in the negative regulation of IP-10 and chemokine production, and interleukin-27 mediated signalling pathway with fold enrichment greater than 50.

In the European genetic model of stroke, the genes associated with latent factor two (F2) did not yield many hits. They are primarily enriched in hippocampal synaptogenesis and neurogenesis, and adipogenesis. In the East Asian genetic model of stroke, the genes associated with latent factor one (F2) are primarily enriched in regulation of insulin secretion, protein transport and metabolism pathways.

The genes associated with each factor were analysed in Open Targets platform [34] to investigate if known drugs can be used as candidates against our latent factors. Multiple hits were obtained for each factor. The drug that can be used against a gene associated with the latent factor, its mode of action and the current diseases it is being used for treatment or clinical trial is shown in table 5 for the European model and in table 6 for the East Asian model.

**Table 5.**
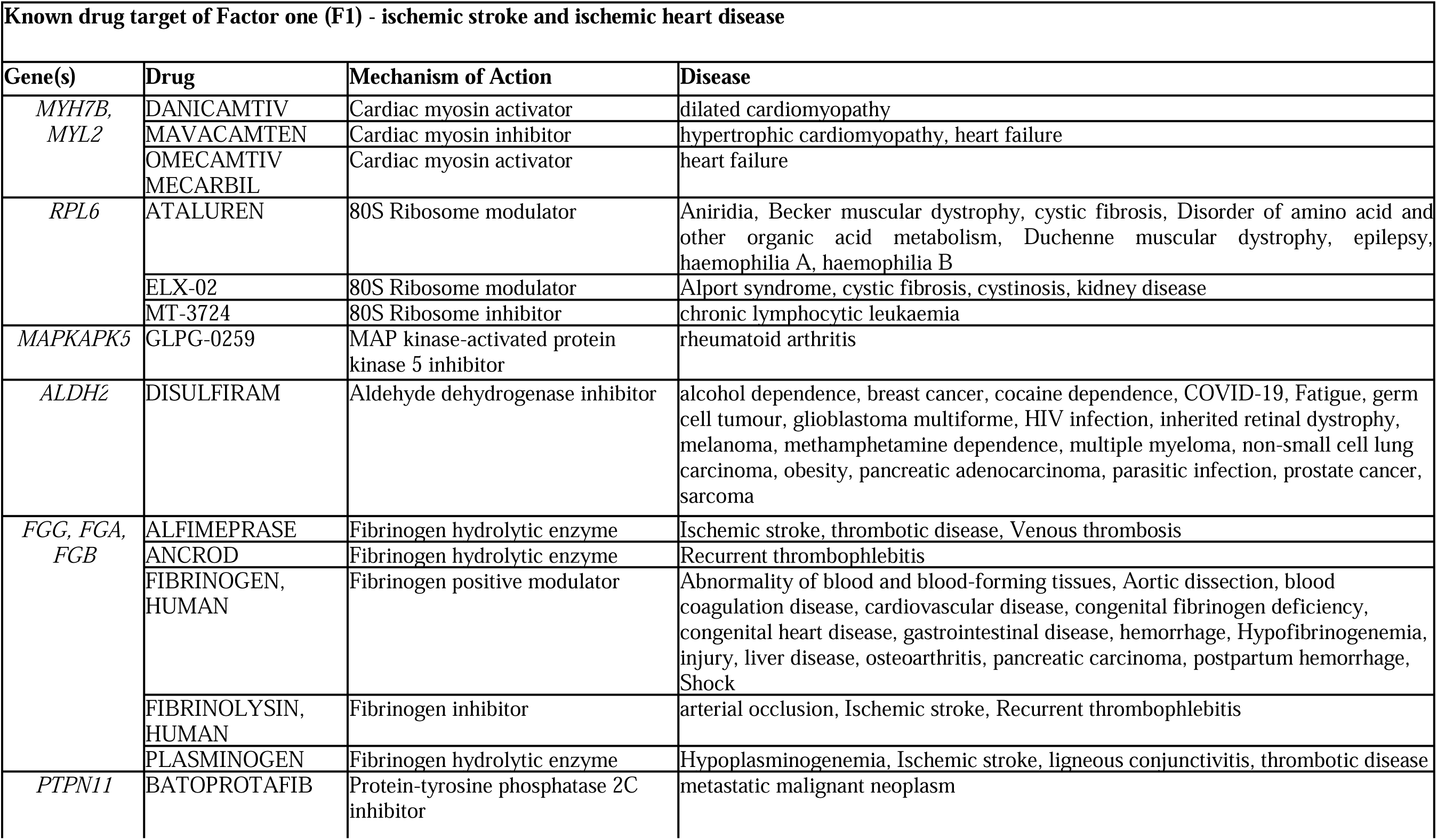
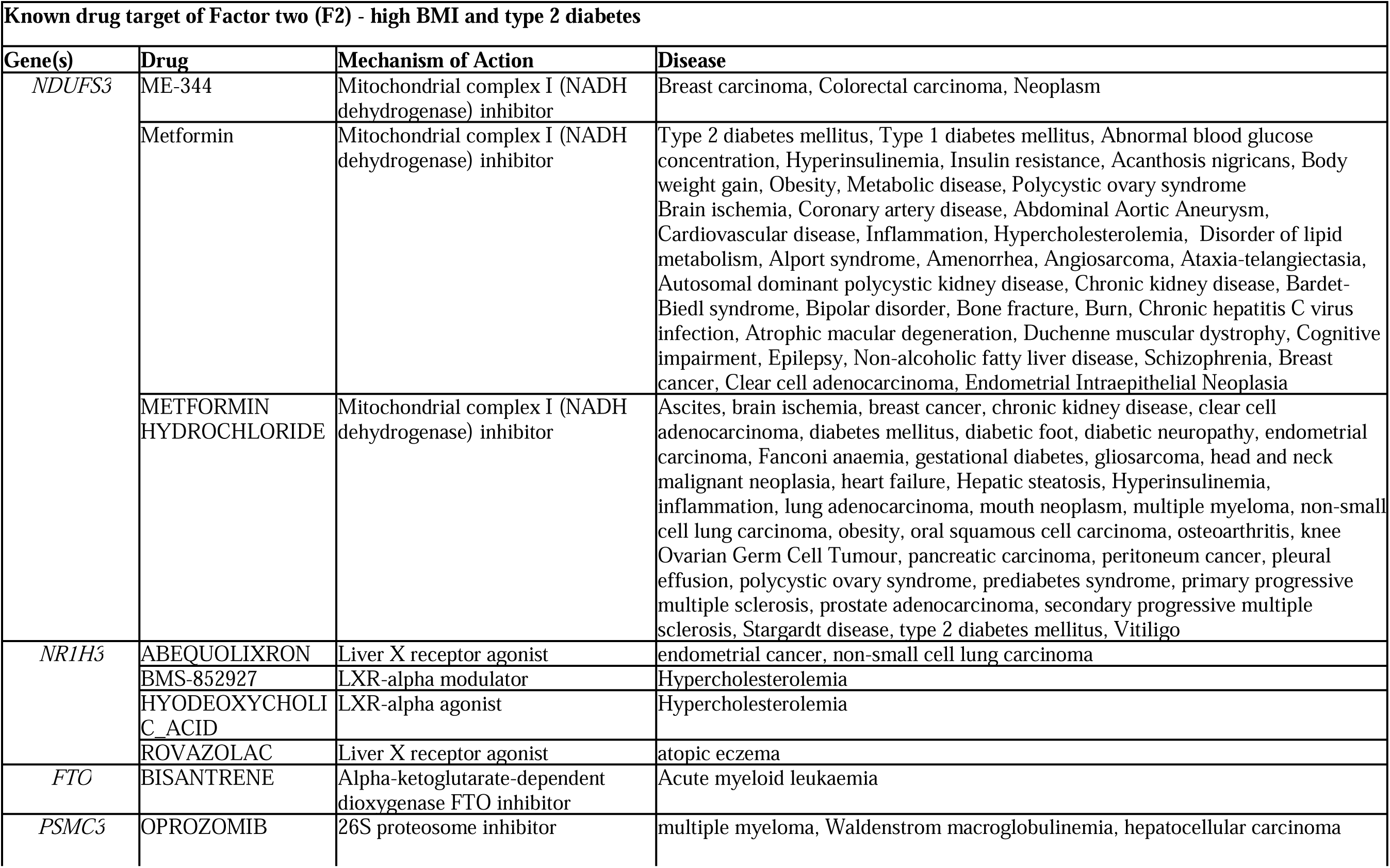
Drug target genes associated with latent factors in the European population. The table shows the genes associated with the latent factors in the European genetic model of stroke, which are known drug targets for various drugs in different phases of clinical trials, along with their model of action and the disease the drugs are used in.

**Table 6.**
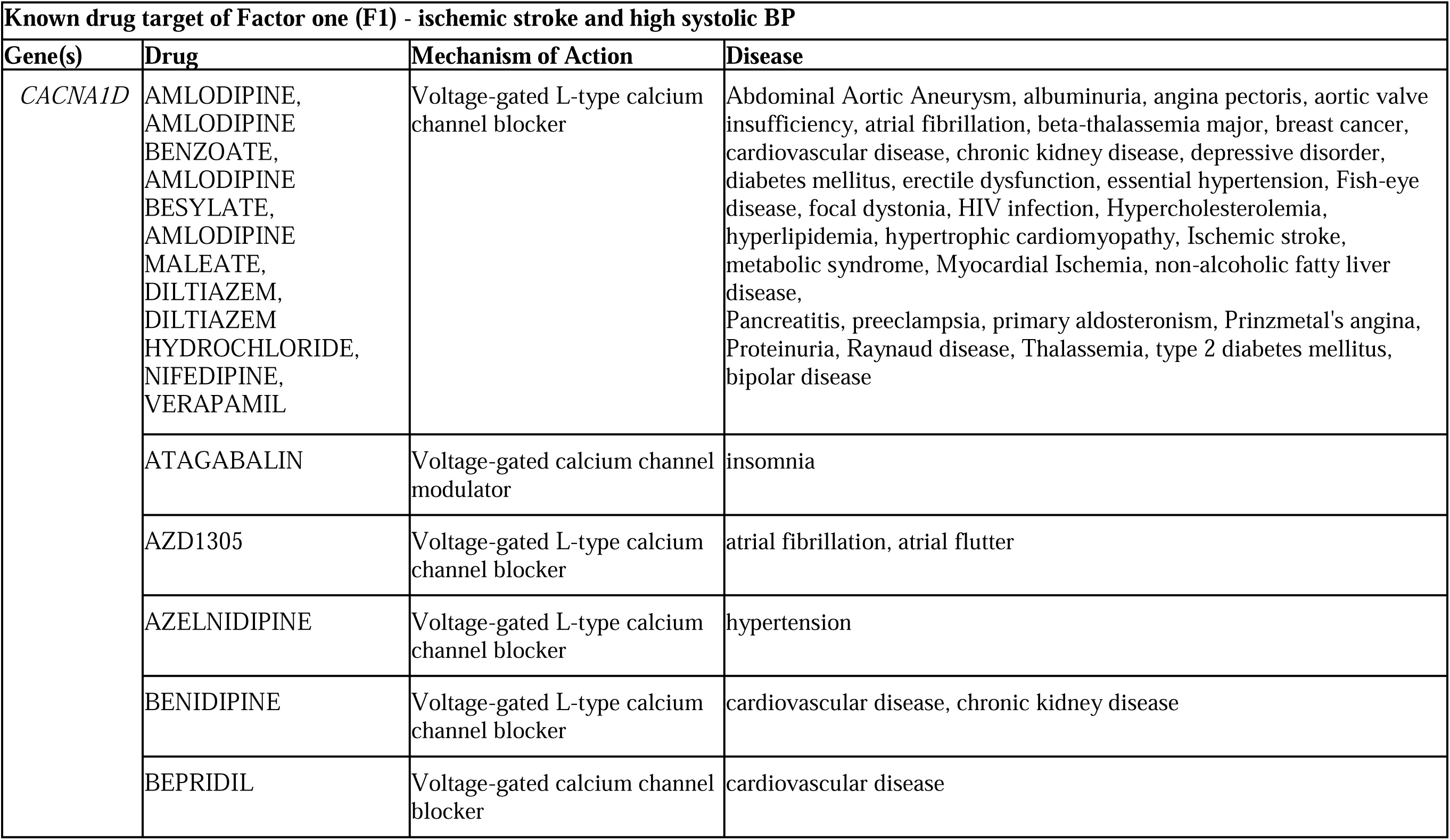
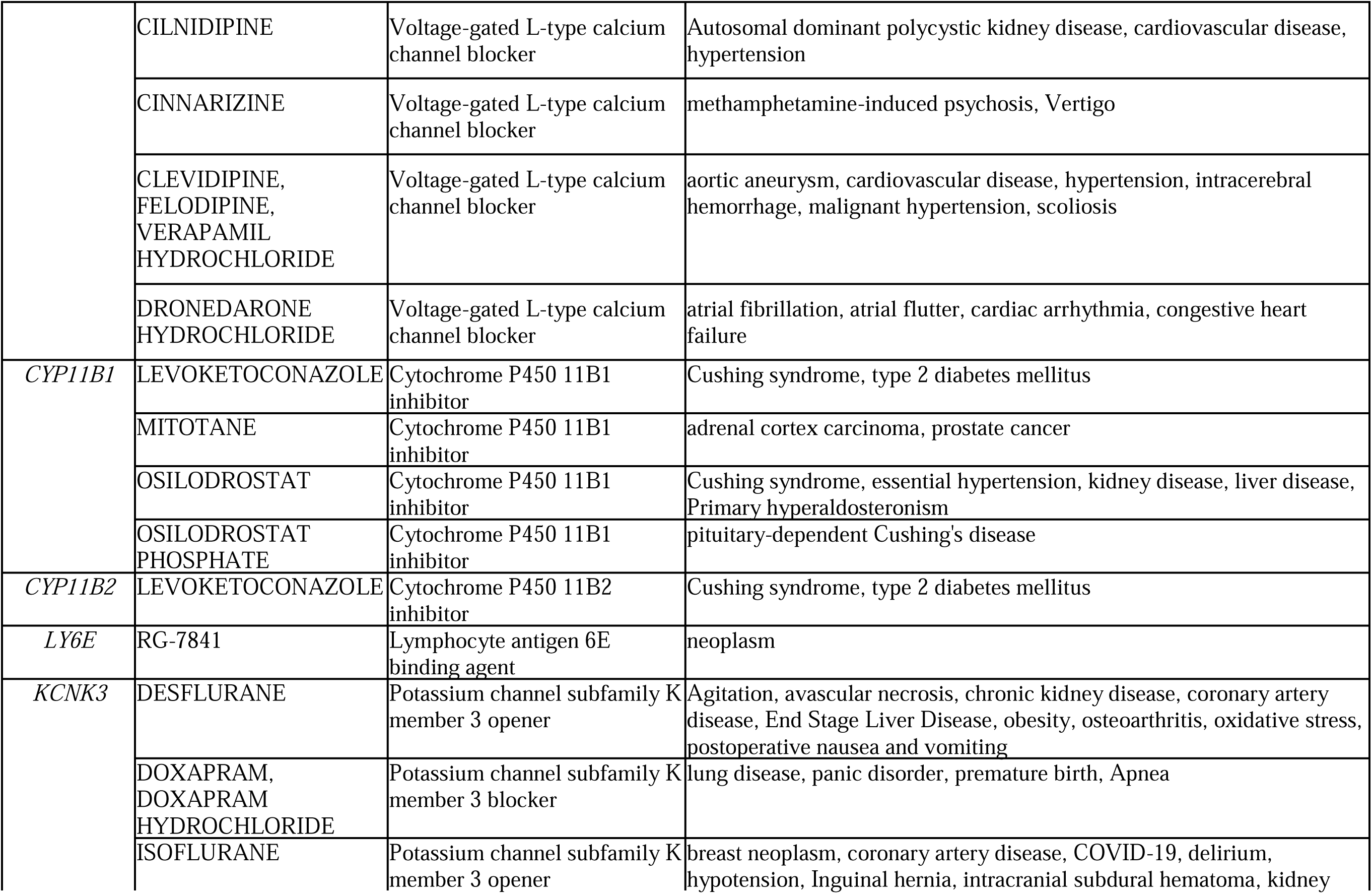
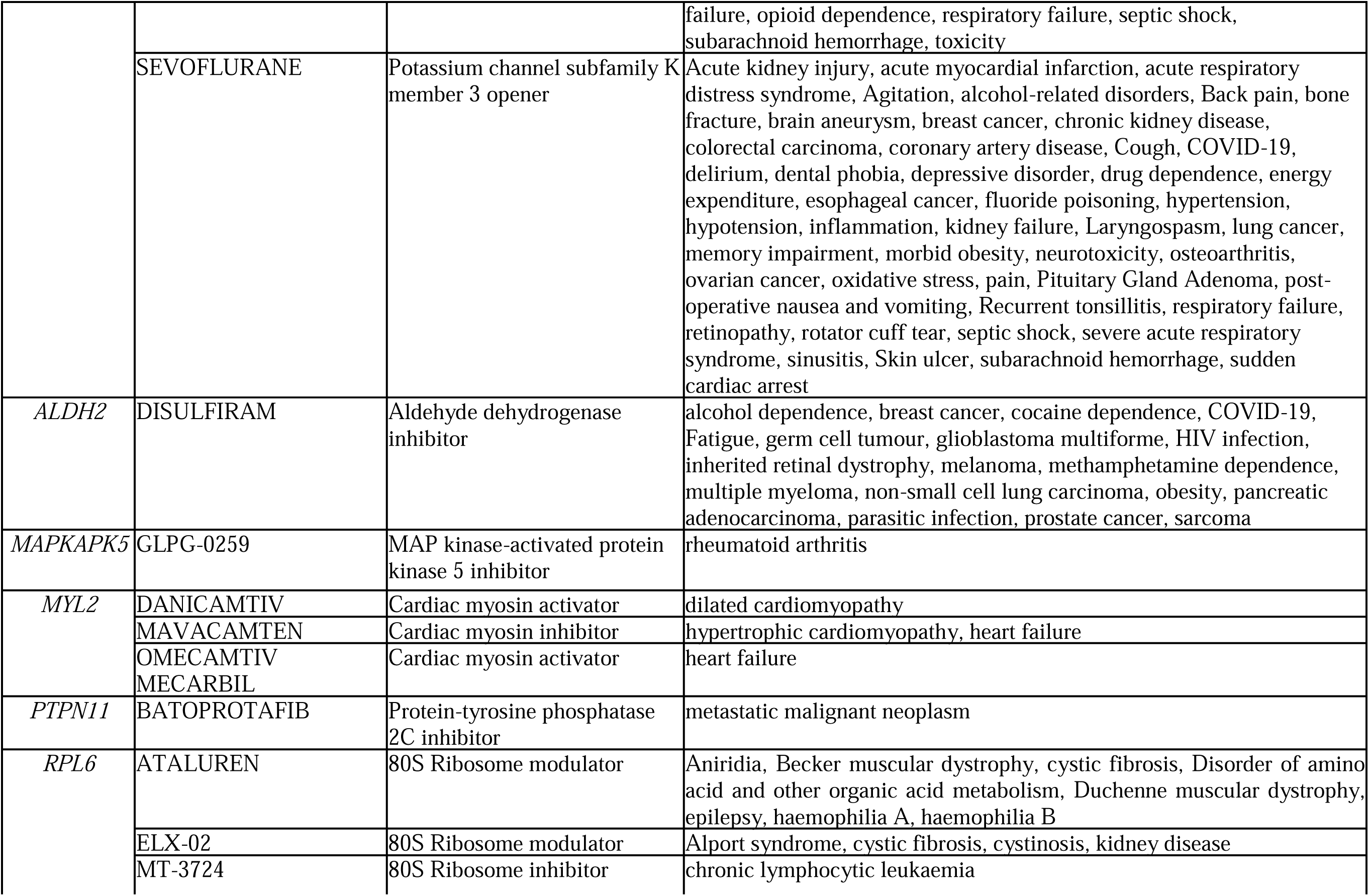
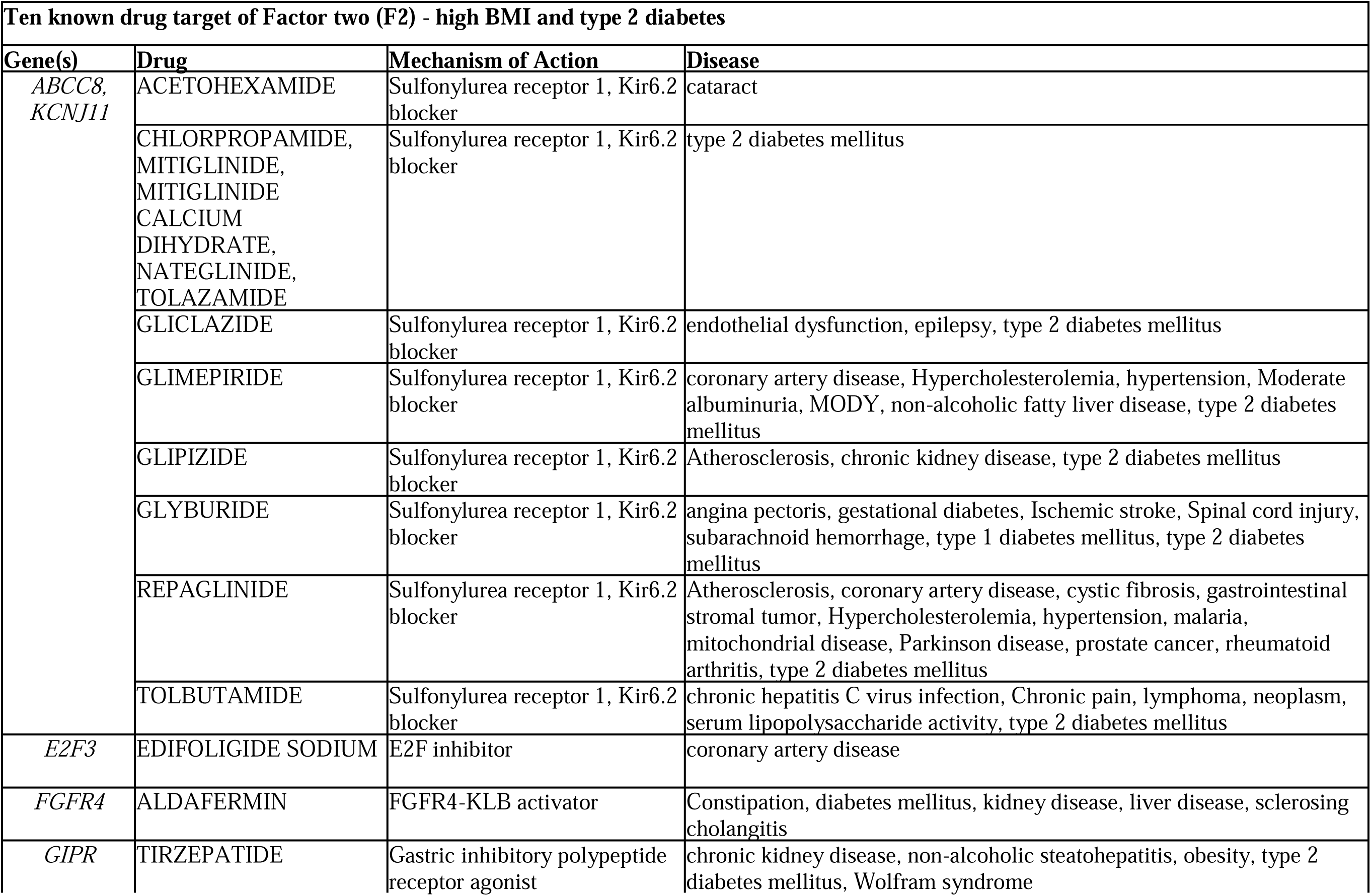
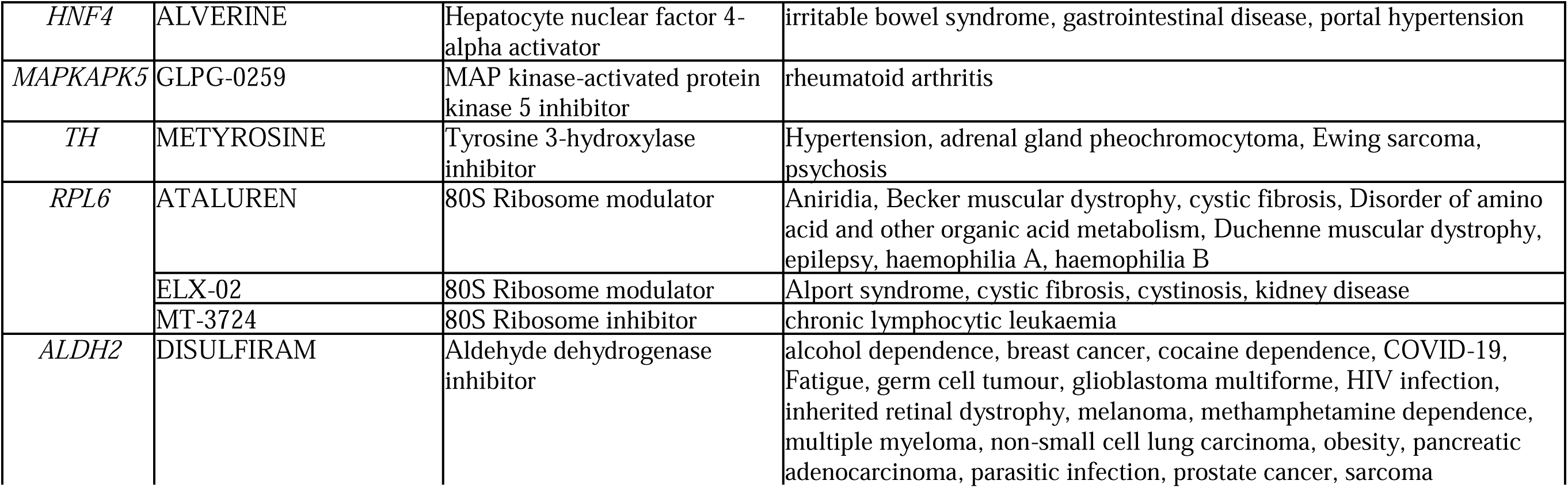
Drug target genes associated with latent factors in the East Asian population. The table shows the genes associated with the latent factors in the East Asian genetic model of stroke, which are known drug targets for various drugs in different phases of clinical trials, along with their model of action and the disease the drugs are used in.

Genomic structural equation modelling is n emerging trend in dissecting the complexities of various diseases, but it has never been investigated from comorbidities point of view. To our knowledge, this is the first study that examined the shared genetic architecture between stroke and its comorbid conditions using genomic structural equation modelling. Stroke and its comorbid conditions were found to be highly correlated genetically as seen in the LDSC genetic correlation analysis. In the European population, the highest genetic correlation is seen between ischemic stroke and ischemic heart disease, followed by ischemic stroke with type 2 diabetes and high systolic BP. In the East Asian population, the highest genetic correlation is seen between type1 and type 2 diabetes, followed closely by type 1 diabetes and chronic kidney disease.

The GWAS heritability of individual traits was observed to vary drastically between the two populations. The highest heritability in the European population was observed for type 1 diabetes, however, it had one of the lowest heritability in East Asia. Interestingly, in the European population, all metabolic diseases have higher heritability than stroke, ischemic heart disease and high systolic BP. On the other hand, the opposite pattern was observed in the East Asian population, where stroke, ischemic heart disease and systolic BP had higher heritability than high LDL cholesterol, type 1 diabetes and chronic kidney disease. This provides evidence to an earlier study where metabolic risks were reported to be higher in Europeans while high systolic blood pressure was observed as the major contributor of risk in East Asians in compared to global average [1]. The differences observed in the epidemiological data are often attributed to differences in the socio-economic parameters [5]. This work presents evidence that the genetic differences among the populations as demonstrated can be the foundation on which the differences in the burden of stroke can be evaluated.

The genetic models can be critical in defining and identifying the latent factors in complex disease conditions, which are often influenced by comorbid factors and sometime even the syndemic drivers. In this direction the study established ethnic-specific genetic models defining the relationships between stroke and its comorbid diseases in the European and East Asian population. In the genetic models for both the populations, two latent factors were identified for stroke and its comorbidity using nearly 12 million SNPs from the European population and 28 million SNPs from East Asia.

In the European model, ischemic stroke and ischemic heart disease loads onto the first genetic latent factor, F1, whereas, in the East Asian model, ischemic stroke and high systolic blood pressure load onto the first factor F1. The second factor in both the models have high BMI and type 2 diabetes loading on to it. From the pathway analysis, genes associated with factor one in both models were involved in coagulation and inflammation related pathways indicating that our factor one, F1, is a possible genetic factor related to inflammation (Figure 7). In both models, high BMI and type 2 diabetes loads onto F2. Thus, the genes being involved in insulin secretion and adipogenesis pathways verifies that our factor two, F2, is a possible genetic metabolic factor (Figure 7). However, the drastic differences in the set of pathways enriched is a clear indicator that latent factors, F1 and F2, in both the models act through entirely different modes in the two populations.

**Figure 7.**
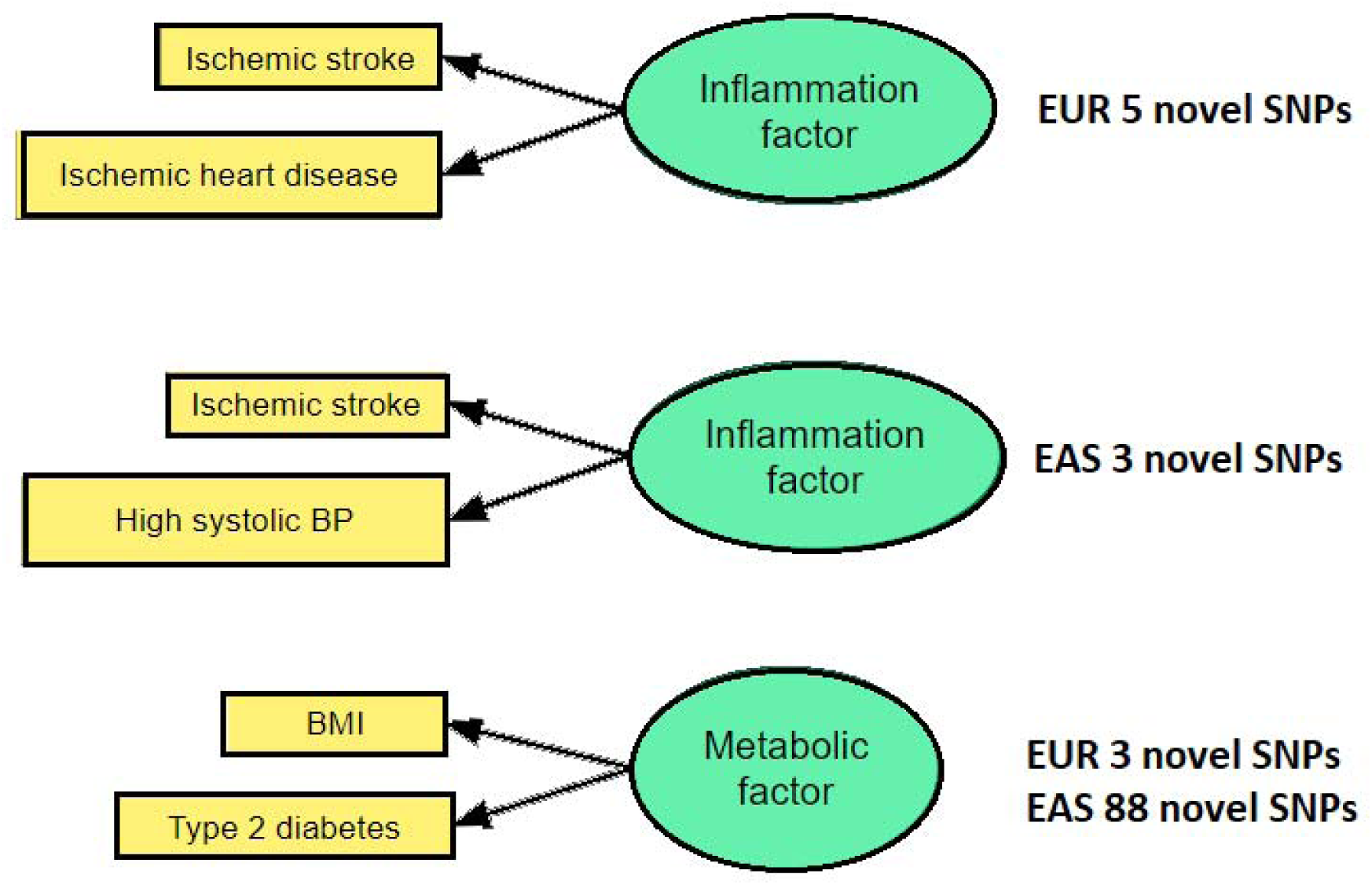
Latent genetic factors and novel SNPs identified for stroke and its comorbidities. The pathways analysis indicates that one latent factor could be an inflammation factor and the other a metabolic factor. The number of novel SNPs identified in the European (EUR) and East Asian (EAS) population for each factor is indicated.

The blood clot formation pathway is enriched for the inflammation factor in the European model. The activation of a cascade of proteins known as clotting factors initiates the coagulation cascade, a sequence of events that leads to the creation of a blood clot following damage [6]. Thus, this indicates that the inflammation factor is mainly acting through the coagulation cascade in the European population. Right after stroke, the inflammation pathway plays a critical role in stroke induced neurodegeneration [7]. The interferon signalling increases the production of pro-inflammatory chemokine IP-10, which in turn drives T helper cells 1 (Th1) response to the site of injury for repair [8]. The negative regulation of IP-10 production is the most enriched pathway for the inflammation factor in East Asia. Decreased IP-10 production has been shown to decrease the infiltration of Th1 cells to the site of injury [9]. Thus, this could be one pathway through which stroke outcomes are affected negatively in East Asia. *ALDH2* 504Lys allele has been reported to be associated with high BMI, increased tolerance of alcohol, high SBP, HDL (high-density lipoprotein), and decreased low density lipoprotein and cardiovascular risk in East Asians [10,11]. This could be the consequence of cross-talk between the two latent factors in East Asia, and thus, again evidence for the validity of our model.

Both the inflammation factor and metabolic factor are genetically defined. In the European population, a total of 11 genome-wide significant and independent SNPs were identified for the inflammation factor, out of which five are novel findings for either ischemic stroke or ischemic heart disease, as these SNPs were not previously identified in any univariate GWAS for the contributing diseases. A total of 14 genome-wide significant and independent SNPs were identified for the metabolic factor, out of which three are novel and have not been reported in GWAS Catalog for either high BMI or type 2 diabetes in any population. Thus, in total, eight new loci are being reported for ischemic stroke and its comorbid conditions, ischemic heart disease, high BMI and type 2 diabetes in the European population. In the East Asian population, a total of 19 genome-wide significant and independent SNPs were identified for the inflammation factor, out of which three are novel and have not been reported in GWAS Catalog for either ischemic stroke or high systolic blood pressure or hypertension. A total of 150 genome-wide significant and independent SNPs were identified for the metabolic factor, out of which 88 are novel and have not been reported in GWAS Catalog for either high BMI or type 2 diabetes. Thus, in total, 91 new loci are being reported for ischemic stroke and its comorbid conditions, high systolic blood pressure, high BMI and type 2 diabetes in the European population. Thus, in total, 99 new loci have been reported for ischemic stroke and its comorbid conditions in the European and East Asian population collectively.

Some SNPs associated with a latent factor, have also been reported for traits in the other latent factor. For instance, the SNP rs11642015 in the gene *FTO* is significantly associated with the metabolic latent factor in East Asia population. This SNP has been reported for traits body mass index [12] and type 2 diabetes [13], as well as for systolic blood pressure [14] in the East Asian population. This cross-trait association is a validation of the genetic model proposed here. It is due to the genetic correlation of these traits through the latent factors that the SNP is seen associated in all three traits.

As our latent factors are upstream to our diseases, these are new potential drug targets, and points of intervention for these diseases. The genes associated with our latent factors have been observed as targets of multiple drugs designed for multiple diseases. Here too, drugs associated with diseases of one factor, are also being used/ tested for a disease from the other factor. This provides evidence for the genetic model proposed that it represents a huge opportunity for drug repurposing among the traits. The association of these variants with the latent factor, and not with the individual diseases is the crucial reason why they had not been discovered in any of the genome wide studies for these diseases. Genomic structural equation modelling (Genomic SEM) has been widely used to study shared genetic architecture of various disorders, like psychiatric conditions [2,15], neurodegenerative disorders [16], chronic pain conditions [17].

It is evident from the genetic evidence presented in this research that there is an ethnic and comorbid specific genetic correlation between stroke and its comorbidities, which shows show a distinct genetic architecture in different populations that possibly does not reflect a clear socio-economic distinction. Though the models look similar, sharing genes among them, the underlying shared genetic architecture is unique to each population, indicating the different treatment modalities that must be employed to minimise adverse reactions. The findings of this research will enable the healthcare industry to reduce the burden of stroke by being aware of the genetic stratification of comorbid conditions, rather than socio-economic stratification in the different populations, while designing prevention programs, creating health policies, collecting public health data and during therapeutic assessments of stroke.

## MATERIALS AND METHODS

### GWAS summary statistics and reference datasets

For identifying the latent factors of stroke and its comorbid conditions, we employed the genomic SEM [2] methodology that uses as input GWAS summary statistics (sumstats). Summary statistics were obtained from GWAS Catalog, [https://www.ebi.ac.uk/gwas/home] [18], during the period January 2023 to November 2023 (Table 7, Supplementary table S1).

**Table 7.**
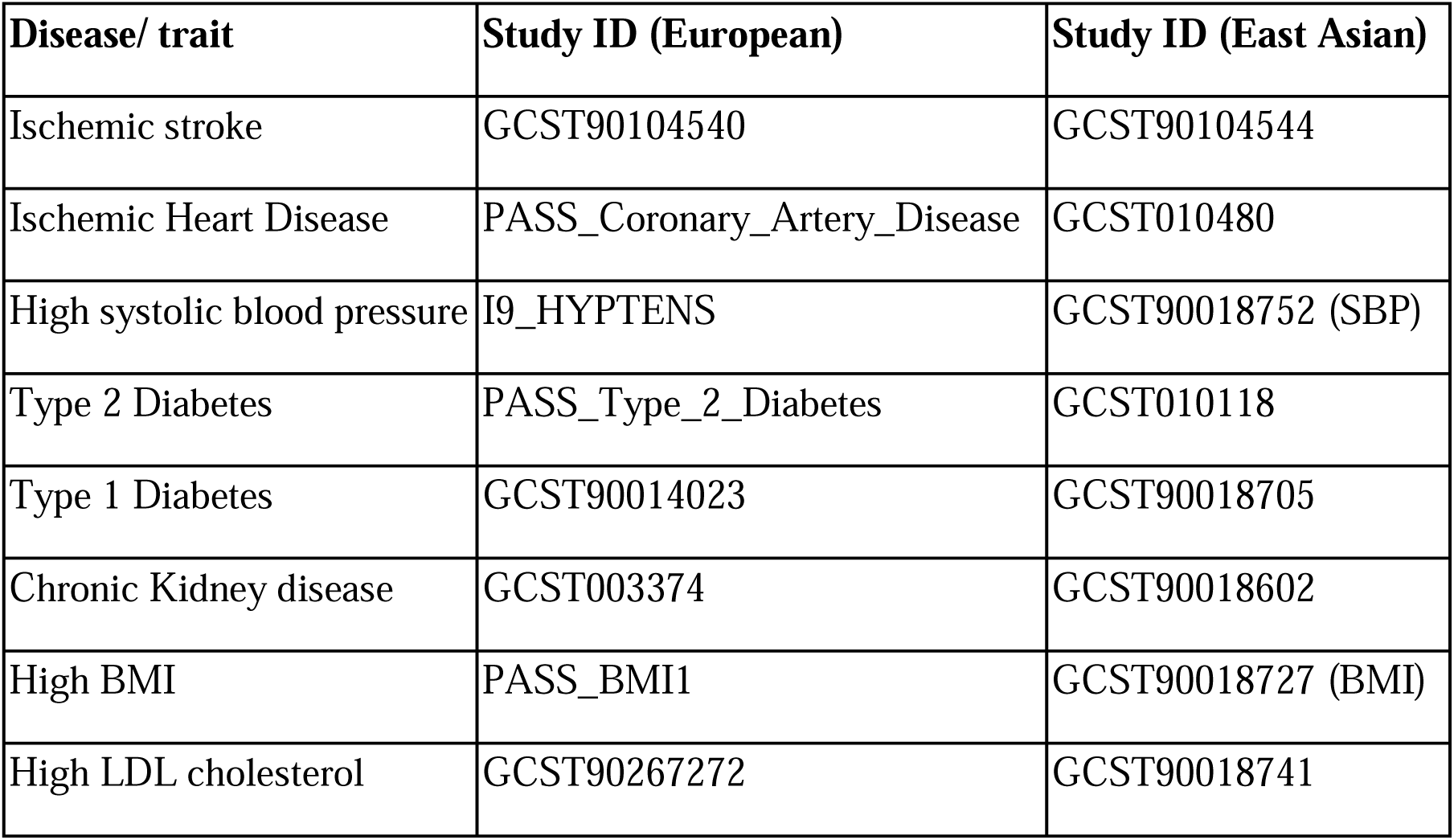
GWAS studies of stroke and its comorbid conditions in European and East Asian populations. The table shows the GWAS study ID of summary statistics downloaded for stroke and its comorbid conditions in the European and East Asian populations.

| Disease/ trait | Study ID (European) | Study ID (East Asian) |
| --- | --- | --- |
| Ischemic stroke | GCST90104540 | GCST90104544 |
| Ischemic Heart Disease | PASS_Coronary_Artery_Disease | GCST010480 |
| High systolic blood pressure | I9_HYPTENS | GCST90018752 (SBP) |
| Type 2 Diabetes | PASS_Type_2_Diabetes | GCST010118 |
| Type 1 Diabetes | GCST90014023 | GCST90018705 |
| Chronic Kidney disease | GCST003374 | GCST90018602 |
| High BMI | PASS_BMI1 | GCST90018727 (BMI) |
| High LDL cholesterol | GCST90267272 | GCST90018741 |

We chose the summary statistics of stroke and its comorbid conditions from the same ancestry. We chose European and East Asian ancestry only, as the GWAS data for stroke and all its comorbid conditions were available only for these ancestries. As the available GWAS summary statistics for ischemic heart disease, high systolic BP, type 2 diabetes, and high BMI for European ancestry failed the pre-processing steps, summary statistics for these diseases/traits were downloaded in October 2023 from the Publicly available summary statistics (PASS_*; https://alkesgroup.broadinstitute.org/sumstats_formatted/) and UK Biobank datasets (https://broad-ukb-sumstats-us-east-1.s3.amazonaws.com/round2/additive-tsvs/I9_HYPTENS.gwas.imputed_v3.both_sexes.tsv.bgz) maintained by Alkes Price group at the Department of Epidemiology at the Harvard School of Public Health. For high systolic blood pressure and high BMI, there were no suitable summary statistics available for the East Asian population, hence summary statistics for systolic blood pressure and normal BMI were used.

Reference panel datasets used were the HapMap3 reference panel [19] and 1000 Genomes Project [20]. Only SNPs with minor allele frequency greater than 0.005 were included, and the sample data from the same ancestry were used. HapMap 3 data and 1000 Genome SNP list for the European population are made available by the developers of Genomic SEM at https://utexas.app.box.com/s/vkd36n197m8klbaio3yzoxsee6sxo11v, and were downloaded in June 2023. For the East Asian population, the list of SNPs was extracted from the 1000 Genomes data using PLINK 1.9 [21]. Pre-computed LD scores for European population, were available at https://utexas.app.box.com/s/vkd36n197m8klbaio3yzoxsee6sxo11v, and downloaded in June 2023, while the pre-computed LD scores for East Asian population, were available at https://zenodo.org/records/7768714/files/1000G_Phase3_EAS_baselineLD_v2.2_ldscores.tgz, and downloaded in November 2023.

### Pre-processing of summary statistics

SNPs in the summary statistics are filtered to match those present in the HapMap 3 and the 1000 Genomes reference panel. This is to ensure uniformity among samples, avoid ambiguous SNPs and also, to reduce bias in the calculation of genetic correlations. The pre-processing was done using the ‘*munge*’ function in the library *genomicSEM* [2] in R Statistical Software (version 4.1.2) [22] using custom scripts adapted from https://github.com/SoskicLab/aid_sharing/tree/main. The PASS_* sumstats files from Alkes group were already pre-processed. The munging process also removes SNPs with missing values, SNPs whose alleles do not match the alleles in the reference panel, and SNPs with minor allele frequency less than 0.01. The final number of SNPs in each summary statistics after munging is shown in Table 8

**Table 8.** The number of SNPs in GWAS summary statistics before and after munging (pre-processing). The table shows the number of SNPs present in the summary statistics of stroke and its comorbid conditions before and after the munging process in the European and East Asian datasets.

| **European Summary Statistics** |  |  |
| --- | --- | --- |
| **Disease/ trait** | **Total SNPs before munge** | **Total SNPs after munge** |
| Ischemic stroke | 7,300,717 | 1,154,619 |
| Ischemic Heart Disease | 925,223 | 924,258 |
| High systolic blood pressure | 8,975,933 | 802,005 |
| Type 2 Diabetes | 968,509 | 967,784 |
| Type 1 Diabetes | 59,696,292 | 1,177,810 |
| Chronic Kidney disease | 2,134,391 | 963,580 |
| High BMI | 1,026,920 | 1,020,822 |
| High LDL cholesterol | 7,107,248 | 1,139,586 |
| **Total SNPs** | **88,135,233** | **8,150,464** |
| **East Asian Summary Statistics** |  |  |
| **Disease/ trait** | **Total SNPs before munge** | **Total SNPs after munge** |
| Ischemic stroke | 6,733,915 | 1,040,752 |
| Ischemic Heart Disease | 5,968,989 | 1,010,927 |
| High systolic blood pressure | 7,138,203 | 1,026,825 |
| Type 2 Diabetes | 7,970,685 | 1,055,287 |
| Type 1 Diabetes | 7,136,430 | 1,026,786 |
| Chronic Kidney disease | 7,136,868 | 1,026,835 |
| High BMI | 7,138,204 | 1,026,827 |
| High LDL cholesterol | 7,137,996 | 1,026,840 |
| **Total SNPs** | **56,361,290** | **8,241,079** |

### Genetic correlation between stroke and its comorbid conditions

Genetic correlation between stroke and its comorbid conditions is calculated using the *‘ldsc’* function in the library *genomicSEM* [2] in R Statistical Software (version 4.1.2) [22]. The function uses LD Score regression [23,24] to calculate the genetic covariance matrix and sample covariance matrix between the traits. This step requires the sample prevalence and population prevalence of each disease. The sample prevalence was calculated from the sample as case/ (case + control) in case of case-control studies or was set to 0.5, if effective population size was available or was set to NA if the trait was a continuous trait. Population prevalence of each disease was obtained from the previous study [1]. The genetic correlation estimated between stroke and its comorbid conditions were plotted using the *corrplot* library [25] in R Statistical software (version 4.1.2) [22]. Summary statistics with a heritability z-score (LDSC heritability score h^2^/ Standard error) of less than five were removed from further analysis, as these would not be able to produce robust genetic correlations. Hence, in the European population, high systolic blood pressure (h^2^z = 2.4) and chronic kidney disease (h^2^z= 3.06) were removed. In the East Asian population, high LDL cholesterol (h^2^z = 4.75), type 1 diabetes (h^2^z = 1.82) and chronic kidney disease (h^2^z = 1.47) were removed.

### Structural equation modelling of stroke and its comorbid conditions

Exploratory Factor Analysis (EFA), to estimate the number of latent factors for stroke and its comorbid conditions in both the European and East Asian population, was done using the ‘promax’ rotation in the *factanal* function in package *stats* in R Statistical software (version 4.1.2) [22]. The scree plot indicating the number of factors was generated using the library *nFactors* [26] in R Statistical software, shown in Supplementary figure S1.

A structural model with hypothesised relationships between our variables, stroke and its comorbid conditions, and one or more latent factors was defined based on the genetic correlation among them derived from the summary statistics and the results of EFA. Confirmatory factor analysis (CFA) was done on structural models defined using the function *usermodel* of *genomicSEM* library in R Statistical software to test how these models fit the data. Though a 2-factor model was suggested for both European and East Asian data in the EFA, we also tested 1-factor, and 3-factor models for completeness. The list of all models tested and their model fit indices are shown in Supplementary table S3. The estimation method used was the Diagonally Weighted Least Square (DWLS), which is the default in usermodel function. A path model to visualise the model and the estimated coefficients, called factor loadings, computed in CFA was plotted using libraries *semPlot* [27] and *lavaan* [28] in R Statistical software (version 4.1.2) [22]. Only models with all factor loadings greater than 0.3 were considered for further analysis. Model fit was evaluated using χ^2^ statistic, comparative fit index [CFI] [29] and standardised root mean square residual (SRMR) [30–33]. A model was considered to be a good fit, if the following criteria were fulfilled - a low χ^2^ statistic, a non-significant p-value, a CFI of 0.95 or greater and an SRMR of 0.05 or lower.

### Common Factor GWAS

Genomic SEM [2] was used to perform a genome wide association study analysis (GWAS) on the selected structural equation model to identify variants that show significant effect on the latent factors, using the function userGWAS in genomic SEM package. Manhattan plots of the summary statistics for each factor were plotted using the *qqman* package [34] in R Statistical software (version 4.1.2) [22].

### Functional Annotation

Summary statistics for each latent factor identified were functionally annotated using FUMA v1.5.2 [35]. Significant single nucleotide polymorphisms (SNPs) with p-value ≤ 5×10^-8^ that were independent (LD measure r^2^ < 0.1 with all other SNPs) were identified as lead SNPs using the *SNP2GENE* function in FUMA. The traits reported associated with each lead SNP was obtained from GWAS Catalog [36] (November 2023). SNPs that did not report a trait present in our model were reported to be novel loci being reported.

Lead SNPs that are physically overlapping with each other or dependent on each other (LD blocks less than 250kb apart) were combined as one genomic risk locus. Thus, a genomic risk locus may contain more than one lead SNP. To validate that the SNPs in a genomic risk locus estimated in the model are not heterogeneous and act only through the model estimated, Q_SNP_ heterogeneity test was done.

SNPs in linkage disequilibrium (LD measure r2 ≥ 0.6) with the lead SNPs and present in the 1000 Genomes reference panel for the respective population (European or East Asian) were identified as candidate SNPs and were selected for further analysis. FUMA maps the candidate SNPs to a nearest protein-coding gene by positional mapping, based on the physical distance of the SNP to the gene (< 10Kb) in the human genome assembly GRCh37. The functional annotation and consequence of all candidate SNPs were done using default parameters. Known functional annotations from Combined Annotation Dependent Depletion [CADD] [37], ANNOVAR [38], and RegulomeDB [39] were used to predict pathogenicity of the variant, functional consequences of the variant and predict regulatory functions, respectively. FUMA also mapped the SNP to a gene (max 1Mb distance) that shows an association between the SNP with the expression level of the gene based on expression quantitative trait loci (eQTLs). The SNPs were mapped to the Brain and Heart tissue types in GTEx v8. Only significant SNP-gene pairs were considered (FDR < 0.05). Ensembl Release 104 [40] was used for all gene mappings, and the major histocompatibility complex region was excluded. The alleles from GWAS summary statistics were aligned with the allele in eQTLs. If the z-score of the allele is positive, then the tested allele increases gene expression, and if negative, it decreases gene expression. On aligning the GWAS alleles with eQTL alleles, if both the alleles are same, then the direction of the GWAS allele is considered same as the eQTL allele, i.e. if positive, then GWAS allele also increases gene expression. If the alleles are not the same, then the direction is considered opposite. Hence, a positive sign (“+”) indicates that the GWAS allele increases gene expression and a negative sign (“-”) indicates the GWAS allele decreases gene expression.

### Pathway analysis and drug target analysis

Pathway enrichment analysis and Gene ontology analysis was performed using ShinyGO V0.76 [41], based on Ensembl Release 104 [40] and STRING-db [42]. Pathway analysis was done using multiple pathway databases like KEGG [43], Wiki pathways [44], Reactome [45], PANTHER [46] and GeneOntology [47]. Open Targets Platform [48] was used to identify drug targets for eQTL genes. This website was queried in December 2023.

## Data Availability

All data produced in the present work are contained in the manuscript

## ACKNOWLEDGEMENTS

RS acknowledges the support of Kerala State Council for Science, Technology and Environment, (KSCSTE) for providing the research fellowship. RS and ASN acknowledge the SIUCEB support at the Department of Computational Biology and Bioinformatics, University of Kerala for providing the necessary facilities to carry out the work. MB acknowledges the BRIC-Dept. of Biotechnology for providing intra-mural support to carry out the work.

## AUTHOR CONTRIBUTIONS

RS, ASN and MB conceptualized and designed the workflow, and wrote the manuscript. RS performed the GSEM analysis. RS, ASN and MB wrote the initial draft and the final draft. ASN and MB provided overall guidance and direction.

## Supplementary Figures and Tables

### Supplementary Tables

**Supplementary Table S1. Details of summary statistics used.**

**Supplementary Table S2. LD Score regression estimates of stroke and its comorbid conditions**

**Supplementary Table S3. CFA model fit indices of one, two factor, three factor models in European and East Asian populations.** The table shows the multiple fit indices, chi square, p-value, AIC, CFI and SRMR for the different models tested in European and East Asian populations. The model passing all indices are highlighted in bold.

**Supplementary Table S4. Overlapping genomic risk loci for latent factor F2 between Europe and East Asia.** The table shows the overlapping risk regions between Europe and East Asia for latent factor F2.

### Supplementary Figures

**Supplementary Figure S1. Scree plot from exploratory factor analysis indicating optimal number of factors for (A) European and (B) East Asian summary statistics.** The different methods indicate that both the European and East Asian datasets optimally have two factors each.

**Supplementary Figure S2A. Confirmatory factor analysis models of stroke and its comorbid conditions with one factor, two factor and three factors in the European population.** The models tested in confirmatory factor analysis with one, two and three latent factors among stroke and its comorbid conditions with factors loadings and correlations between them.

**Supplementary Figure S2B. Confirmatory factor analysis models of stroke and its comorbid conditions with one factor, two factor and three factors in the East Asian population.** The models tested in confirmatory factor analysis with one, two and three latent factors among stroke and its comorbid conditions with factors loadings and correlations between them.

**Supplementary Figure S3. Secondary genetic models proposed for stroke and its comorbid conditions in the European population.** The second proposed genetic models for stroke and its comorbid conditions have two latent factors. Ischemic stroke (is) and ischemic heart disease (ihd) loads onto the first factor F1, and high body mass index (bmi), type 2 diabetes (t2d) as well as F1 loads onto the second factor F2. Both factors, F1 and F2, have path coefficients to each variable greater than 0.3. Factor 1 also loads on factor 2 with a path coefficient of 0.56. The residuals of the variables are shown in circular self-paths.

**Supplementary Figure S4. Secondary genetic models proposed for stroke and its comorbid conditions in the East Asian population.** The second proposed genetic models for stroke and its comorbid conditions have two latent factors. Ischemic stroke (is) and high systolic blood pressure (hyp) loads onto the first factor F1, and high body mass index (bmi), type 2 diabetes (t2d) as well as F1 loads onto the second factor F2. Both factors, F1 and F2, have path coefficients to each variable greater than 0.3. Factor 1 also loads on factor 2 with a path coefficient of 0.67. The residuals of the variables are shown in circular self-paths.

**Supplementary Figure S5. Manhattan plot of GWAS of two latent factors F1 and F2 of stroke and its comorbid conditions in European population.** The plot shows all the SNPs analysed for each factor F1 and F2 as a dot. The X-axis shows the chromosome location of the SNP, the y-axis shows the negative log of p-value. SNPs above the threshold line of 5e-08 are considered significant.

**Supplementary Figure S6. Manhattan plot of GWAS of two latent factors F1 and F2 of stroke and its comorbid conditions in East Asian population.** The plot shows all the SNPs analysed for each factor F1 and F2 as a dot. The X-axis shows the chromosome location of the SNP, the y-axis shows the negative log of p-value. SNPs above the threshold line of 5e-08 are considered significant.

**Supplementary Figure S7. Q_SNP_ heterogeneity test per genomic risk locus of latent factors F1 and F2 in the European and East Asian models.** The chi-square tests the null hypothesis that the SNPs in the genomic risk loci act through the defined model. Hence, if significant, the SNPs are indeed heterogeneous and the test returns TRUE. If non-significant, the SNPs are acting only through our model, and thus, the test returns FALSE.

**Supplementary Figure S8. The gene location (exonic, downstream and UTR regions) of the SNPs associated with the latent factors.** The histogram displays the proportion of all SNPs in LD with significant SNPs which have corresponding functional annotation from ANNOVAR. Bars are coloured by log2(enrichment) relative to all SNPs selected in the reference panel. EUR F1 and EUR F2 are factors in European population and EAS F1 and EAS F2 are factors in East Asian population.

## REFERENCES

1. Sukumaran Rashmi, Nair Achuthsankar S., Banerjee Moinak (2024) Ethnic and region-specific genetic risk variants of stroke and its comorbid conditions can define the variations in the burden of stroke and its phenotypic traits eLife 13:RP94088.

2. Grotzinger AD, Rhemtulla M, de Vlaming R, Ritchie SJ, Mallard TT, et al. Genomic structural equation modelling provides insights into the multivariate genetic architecture of complex traits. Nat Hum Behav. 2019 May;3(5):513–525.

3. Hoyle RH. Handbook of structural equation modeling. 2012. The Guilford Press. ISBN: 9781462544646.

4. Field A. Discovering Statistics Using IBM SPSS Statistics. 2013. SAGE Publications. ISBN: 9781526445780.

5. GBD 2019 Stroke Collaborators. Global, regional, and national burden of stroke and its risk factors, 1990-2019: a systematic analysis for the Global Burden of Disease Study 2019. Lancet Neurol. 2021 Oct;20(10):795–820.

6. Palta S, Saroa R, Palta A. Overview of the coagulation system. Indian journal of anaesthesia. 2014 Sep 1;58(5):515–23

7. Leonardo CC, Pennypacker KR. The splenic response to ischemic stroke: what have we learned from rodent models? Transl Stroke Res. 2011 Sep;2(3):328–38.

8. Loetscher P, Pellegrino A, Gong JH, Mattioli I, Loetscher M, et al. The ligands of CXC chemokine receptor 3, I-TAC, Mig, and IP10, are natural antagonists for CCR3. J Biol Chem. 2001 Feb 2;276(5):2986-91.

9. Seifert HA, Collier LA, Chapman CB, Benkovic SA, Willing AE, Pennypacker KR. Pro-inflammatory interferon gamma signaling is directly associated with stroke induced neurodegeneration. J Neuroimmune Pharmacol. 2014 Dec;9(5):679–89.

10. Takeuchi F, Isono M, Nabika T, Katsuya T, Sugiyama T, et al. Confirmation of ALDH2 as a Major locus of drinking behavior and of its variants regulating multiple metabolic phenotypes in a Japanese population. Circ J. 2011;75(4):911–8.

11. Xu F, Chen Y, Lv R, Zhang H, Tian H, et al. ALDH2 genetic polymorphism and the risk of type II diabetes mellitus in CAD patients. Hypertens Res. 2010 Jan;33(1):49–55.

12. Akiyama M, Okada Y, Kanai M, Takahashi A, Momozawa Y, et al. Genome-wide association study identifies 112 new loci for body mass index in the Japanese population. Nat Genet. 2017 Oct;49(10):1458–1467.

13. Ishigaki K, Akiyama M, Kanai M, Takahashi A, Kawakami E, et al. Large-scale genome-wide association study in a Japanese population identifies novel susceptibility loci across different diseases. Nat Genet. 2020 Jul;52(7):669–679.

14. Takeuchi F, Akiyama M, Matoba N, Katsuya T, Nakatochi M, et al. Interethnic analyses of blood pressure loci in populations of East Asian and European descent. Nat Commun. 2018 Nov 28;9(1):5052.

15. Morey RA, Zheng Y, Bayly H, Sun D, Garrett ME, et al. Genomic structural equation modeling reveals latent phenotypes in the human cortex with distinct genetic architecture. Transl Psychiatry 2024 October 24; 14, 451.

16. Foote IF, Jacobs BM, Mathlin G, Watson CJ, Bothongo PL, et al. The shared genetic architecture of modifiable risk for Alzheimer’s disease: a genomic structural equation modelling study. Neurobiol Aging. 2022 Sep;117:222–235.

17. Zorina-Lichtenwalter K, Bango CI, Van Oudenhove L, Čeko M, Lindquist MA, et al. Genetic risk shared across 24 chronic pain conditions: identification and characterization with genomic structural equation modeling. Pain. 2023 Oct 1;164(10):2239–2252.

18. Sollis E, Mosaku A, Abid A, Buniello A, Cerezo M, et al. The NHGRI-EBI GWAS Catalog: knowledgebase and deposition resource. Nucleic Acids Res. 2023 Jan 6;51(D1):D977–D985.

19. International HapMap Consortium. The International HapMap Project. Nature. 2003 Dec 18;426(6968):789-96.

20. 1000 Genomes Project Consortium; Auton A, Brooks LD, Durbin RM, Garrison EP, et al. A global reference for human genetic variation. Nature. 2015 Oct 1;526(7571):68-74.

21. Chang CC, Chow CC, Tellier LC, Vattikuti S, Purcell SM, et al. Second-generation PLINK: rising to the challenge of larger and richer datasets. Gigascience. 2015 Feb 25;4:7.

22. R Core Team. R: A language and environment for statistical computing. R Foundation for Statistical Computing, Vienna, Austria. 2021. URL: https://www.R-project.org/.

23. Bulik-Sullivan BK, Loh PR, Finucane HK, Ripke S, Yang J, et al. LD Score regression distinguishes confounding from polygenicity in genome-wide association studies. Nat Genet. 2015 Mar;47(3):291–5.

24. Bulik-Sullivan BK, Finucane HK, Anttila V, Gusev A, Day FR, et al. An atlas of genetic correlations across human diseases and traits. Nat Genet. 2015 Nov;47(11):1236–41.

25. Wei T, Simko V. ‘corrplot’: Visualization of a Correlation Matrix, R package version 0.92. 2021. URL: https://github.com/taiyun/corrplot.

26. Raiche G, Magis D. nFactors: Parallel Analysis and Other Non Graphical Solutions to the Cattell Scree Test. R package version 2.4.1.1, 2022

27. Epskamp S. semPlot: Path Diagrams and Visual Analysis of Various SEM Packages Output. R package version 1.1.6, 2022.

28. Rosseel Y. lavaan: An R Package for Structural Equation Modeling. Journal of Statistical Software. 2012;48(2):1-36.

29. Bentler PM. Comparative fit indexes in structural models. Psychol Bull. 1990 Mar;107(2):238–46.

30. Jöreskog, KG, Sörbom D. LISREL 8: Structural equation modeling with the SIMPLIS command language. 1993. Scientific Software International; Lawrence Erlbaum Associates, Inc. ISBN: 0894980335.

31. Hu L, Bentler PM. Cutoff criteria for fit indexes in covariance structure analysis: Conventional criteria versus new alternatives. Structural Equation Modeling: A Multidisciplinary Journal. 1999;6(1), 1–55.

32. Bollen, KA, Long JS. Testing structural equation models. 1993 FEb. SAGE Focus Editions V53, Sage Publications, Inc. ISBN: 9780803945074.

33. Byrne BM. Structural equation modeling with LISREL, PRELIS, and SIMPLIS: Basic concepts, applications, and programming. 1998. Lawrence Erlbaum Associates Publishers. eISBN: 9780203774762.

34. Turner SD. qqman: an R package for visualizing GWAS results using Q-Q and manhattan plots. Journal of Open Source Software. 2018; 3(25):731.

35. Watanabe K, Taskesen E, van Bochoven A, Posthuma D. Functional mapping and annotation of genetic associations with FUMA. Nat Commun. 2017 Nov 28;8(1):1826..

36. Sollis E, Mosaku A, Abid A, Buniello A, Cerezo M, et al. The NHGRI-EBI GWAS Catalog: knowledgebase and deposition resource. Nucleic Acids Res. 2023 Jan 6;51(D1):D977–D985..

37. Rentzsch P, Witten D, Cooper GM, Shendure J, Kircher M. CADD: predicting the deleteriousness of variants throughout the human genome. Nucleic Acids Res. 2019 Jan 8;47(D1):D886–D894.

38. Wang K, Li M, Hakonarson H. ANNOVAR: functional annotation of genetic variants from high-throughput sequencing data. Nucleic Acids Res. 2010 Sep;38(16):e164.

39. Boyle AP, Hong EL, Hariharan M, Cheng Y, Schaub MA, et al. Annotation of functional variation in personal genomes using RegulomeDB. Genome Res. 2012 Sep;22(9):1790–7.

40. Yates A, Akanni W, Amode MR, Barrell D, Billis K, et al. Ensembl 2016. Nucleic Acids Res. 2016 Jan 4;44(D1):D710-6.

41. Ge SX, Jung D, Yao R. ShinyGO: a graphical gene-set enrichment tool for animals and plants. Bioinformatics. 2020 Apr 15;36(8):2628–2629.

42. Szklarczyk D, Gable AL, Nastou KC, Lyon D, Kirsch R, et al. The STRING database in 2021: customizable protein-protein networks, and functional characterization of user-uploaded gene/measurement sets. Nucleic Acids Res. 2021;49(D1):D605–D612.

43. Kanehisa M, Goto S. KEGG: kyoto encyclopedia of genes and genomes. Nucleic Acids Res. 2000 Jan 1;28(1):27–30.

44. Pico AR, Kelder T, van Iersel MP, Hanspers K, Conklin BR, et al. WikiPathways: pathway editing for the people. PLoS Biol. 2008 Jul 22;6(7):e184.

45. Gillespie M, Jassal B, Stephan R, Milacic M, Rothfels K, et al. The reactome pathway knowledgebase 2022. Nucleic Acids Res. 2022 Jan 7;50(D1):D687–D692.

46. Thomas PD, Ebert D, Muruganujan A, Mushayahama T, Albou LP, et al. PANTHER: Making genome-scale phylogenetics accessible to all. Protein Sci. 2022;31(1):8–22.

47. Gene Ontology Consortium. Gene Ontology Consortium: going forward. Nucleic Acids Res. 2015 Jan;43(Database issue):D1049-56.

48. Ochoa D, Hercules A, Carmona M, Suveges D, Baker J, et al. The next-generation Open Targets Platform: reimagined, redesigned, rebuilt. Nucleic Acids Res. 2023 Jan 6;51(D1):D1353–D1359.

