## Supplementary file for "Shared genetic architecture of stroke and its comorbid conditions reveals latent phenotypes that differ among populations: A genomic structural equation modelling approach"

### **Supplementary Information**

### **Supplementary Tables S1-S4**

### **Supplementary Figures S1-S8**

**Supplementary Table S1. Details of summary statistics used.**

| **European datasets** | | | | |
| --- | --- | --- | --- | --- |
| **Trait** | **Sample size**  **(cases + controls)** | **GWAS catalog/ link** | **Genome build** | **Reference** |
| Ischemic Stroke | 1296908  (62100 + 1234808) | <https://www.ebi.ac.uk/gwas/studies/GCST90104540> | build 37 | Mishra et al., 2022 |
| Ischemic Heart Disease (IHD) | 86995  (22233 + 64762) | https://storage.googleapis.com/broad-alkesgroup-public/sumstats_formatted/PASS_Coronary_Artery_Disease.sumstats | build 37 | Schunkert, 2011 |
| High LDL | 269545  (NA) | <https://www.ebi.ac.uk/gwas/studies/GCST90267272> | build 37 | Schoeler T, et al., 2023 |
| High systolic blood pressure (High SBP) | 361194  (359957 +1237) | http://www.nealelab.is/uk-biobank/ | build 37 | Sudlow, et al. 2015 |
| High body mass index (High BMI) | 123865  (NA) | https://storage.googleapis.com/broad-alkesgroup-public/sumstats_formatted/PASS_BMI1.sumstats | build 37 | Speliotes EK, et al., 2010 |
| Chronic Kidney Disease (CKD) | 117165  (12385 + 104780) | <https://www.ebi.ac.uk/gwas/studies/GCST003374> | build 37 | Pattaro C, et al., 2016 |
| Type 1 diabetes (T1D) | 520580  (18942 +501638) | https://www.ebi.ac.uk/gwas/studies/GCST90014023 | build 38 | Chiou J, et al., 2021 |
| Type 2 diabetes (T2D) | 69033  (12171 + 56862) | https://storage.googleapis.com/broad-alkesgroup-public/sumstats_formatted/PASS_Type_2_Diabetes.sumstats | build 37 | Morris AP, et al., 2012 |
| **East Asian datasets** | | | | |
| **Trait** | **Sample size**  **(cases + controls)** | **GWAS catalog** | **Genome build** | **Reference** |
| Ischemic Stroke | 264655  (27413 + 237242) | [https://www.ebi.ac.uk/gwas/studies/](https://www.ebi.ac.uk/gwas/studies/GCST90104540)GCST90104544 | build 37 | Mishra et al., 2022 |
| Ischemic Heart Disease (IHD) | 51442  (15,302 + 36,140) | [https://www.ebi.ac.uk/gwas/studies/GCST](https://www.ebi.ac.uk/gwas/studies/GCST000998)010480 | build 37 | Matsunaga H, et al., 2020 |
| High LDL | 72866  (NA) | [https://www.ebi.ac.uk/gwas/studies/GCST90018741](https://www.ebi.ac.uk/gwas/studies/GCST90267272) | build 37 | Sakaue S, et al., 2021 |
| High systolic blood pressure (High SBP) | 145505  (NA) | [https://www.ebi.ac.uk/gwas/studies/GCST90018752](https://www.ebi.ac.uk/gwas/studies/GCST007228) | build 37 | Sakaue S, et al., 2021 |
| High body mass index (High BMI) | 163835  (NA) | [https://www.ebi.ac.uk/gwas/studies/GCST](https://www.ebi.ac.uk/gwas/studies/GCST001676)90018727 | build 37 | Sakaue S, et al., 2021 |
| Chronic Kidney Disease (CKD) | 176462  (2,117 + 174,345) | [https://www.ebi.ac.uk/gwas/studies/GCST](https://www.ebi.ac.uk/gwas/studies/GCST003374)90018602 | build 37 | Sakaue S, et al., 2021 |
| Type 1 diabetes (T1D) | 133251  (1,219 + 132,032) | https://www.ebi.ac.uk/gwas/studies/GCST90018705 | build 37 | Sakaue S, et al., 2021 |
| Type 2 diabetes (T2D) | 433540  (77,418 + 356,122) | [https://www.ebi.ac.uk/gwas/studies/GCST010118](https://www.ebi.ac.uk/gwas/studies/GCST90006934) | build 37 | Spracklen CN, et al., 2020 |

**Supplementary Table S2. LD Score regression estimates of stroke and its comorbid conditions**

| **LD score regression estimates in European population** | | | | | | | |
| --- | --- | --- | --- | --- | --- | --- | --- |
| **Trait** | **Number of variants** | **h2 (SE)** | **h2 z-score** | **Mean χ2** | **λGC** | **Intercept (SE)** | **Ratio (SE)** |
| Ischemic stroke | 1154619 | 0.0415 (0.0032) | 13 | 1.2282 | 1.1893 | 1.0697 (0.0074) | 0.3057 (0.0323) |
| Ischemic heart disease | 924258 | 0.076 (0.0096) | 7.93 | 1.143 | 1.105 | 1.0214 (0.0092) | 0.149 (0.0643) |
| High LDL cholesterol | 1139586 | 0.0811 (0.009) | 9.01 | 1.5174 | 1.2391 | 1.0633 (0.0361) | 0.1224 (0.0697) |
| High systolic BP | 802005 | 0.0034 (0.0014) | 2.4 | 1.0202 | 1.0218 | 0.9975 (0.0065) | -0.1222 (0.3234) |
| High BMI | 1020822 | 0.1439 (0.0078) | 18.3 | 1.1099 | 1.0378 | 0.7639 (0.0099) | -2.1486 (0.0901) |
| Chronic kidney disease | 963580 | 0.0177 (0.0058) | 3.06 | 1.0624 | 1.0475 | 1.0175 (0.0089) | 0.2807 (0.1423) |
| Type 1 diabetes | 1177810 | 0.2858 (0.0257) | 11.1 | 1.3321 | 1.1999 | 1.0682 (0.0121) | 0.2055 (0.0365) |
| Type 2 diabetes | 967784 | 0.095 (0.0108) | 8.76 | 1.1292 | 1.0956 | 1.012 (0.0086) | 0.0932 (0.0668) |
| **LD score regression estimates in East Asian population** | | | | | | | |
| **Trait** | **Number of variants** | **h2 (SE)** | **h2 z-score** | **Mean χ2** | **λGC** | **Intercept (SE)** | **Ratio (SE)** |
| Ischemic stroke | 1040752 | 0.0423 (0.0063) | 6.71 | 1.1163 | 1.0986 | 1.0332 (0.0076) | 0.2853 (0.0652) |
| Ischemic heart disease | 1010927 | 0.0811 (0.0115) | 7.06 | 1.0951 | 1.0935 | 1.0221 (0.0075) | 0.232 (0.0789) |
| High LDL cholesterol | 1026840 | 0.036 (0.0076) | 4.75 | 1.1548 | 1.0966 | 1.0387 (0.017) | 0.25 (0.11) |
| High systolic BP | 1026825 | 0.0779 (0.0072) | 10.8 | 1.2971 | 1.1999 | 1.0566 (0.0111) | 0.1905 (0.0373) |
| High BMI | 1026827 | 0.1778 (0.0076) | 23.4 | 1.6885 | 1.4921 | 1.0765 (0.0132) | 0.1111 (0.0191) |
| Chronic kidney disease | 1026835 | 0.0036 (0.0025) | 1.47 | 1.0084 | 1.0042 | 0.9946 (0.0067) | -0.6513 (0.803) |
| Type 1 diabetes | 1026786 | 0.0062 (0.0034) | 1.82 | 1.0048 | 1.0012 | 0.9871 (0.0067) | -2.6739 (1.3971) |
| Type 2 diabetes | 1055287 | 0.1514 (0.0083) | 18.2 | 1.6553 | 1.3602 | 1.0026 (0.0182) | 0.004 (0.0278) |

**Supplementary Table S3. CFA model fit indices of one, two factor, three factor models in European and East Asian populations.** The table shows the multiple fit indices, chi square, p-value, AIC, CFI and SRMR for the different models tested in European and East Asian populations. The model passing all indices are highlighted in bold.

| **Model fit indices for models tested in European population** | | | | | | |
| --- | --- | --- | --- | --- | --- | --- |
| **CFA Model (one factor)** | **ChiSq** | **df** | **p-value** | **AIC** | **CFI** | **SRMR** |
| F1 =~ NA*is + ihd + t2d + bmi | 16.25864 | 2 | 0.0002947682 | 32.25864 | 0.9247417 | 0.06215042 |
| F1 =~ NA*is + ihd + t2d + bmi + ldl + t1d | 30.89718 | 9 | 0.0003082971 | 54.89718 | 0.9084415 | 0.0563456 |
| F1 =~ NA*is + ihd + t2d + bmi + ldl | 25.73752 | 5 | 0.0001003266 | 45.73752 | 0.90034 | 0.06115894 |
| F1 =~ NA*is + ihd + t2d + bmi + t1d | 17.07842 | 5 | 0.004353442 | 37.07842 | 0.9396306 | 0.05598006 |
| **CFA Model (two factor)** |  |  |  |  |  |  |
| **F1 =~ NA*is + ihd + F2 =~ NA*bmi + t2d** | **0.01535461** | 1 | **0.9013836** | 18.01535 | **1** | **0.001675339** |
| **F1 =~ NA*is + ihd + F2 =~ NA*bmi + t2d + F1** | **0.01535461** | 1 | **0.9013836** | 18.01535 | **1** | **0.001675339** |
| F1 =~ NA*ihd + t1d + F2 =~ NA*bmi + t2d + is | 19.00253 | 4 | 0.0007850462 | 41.00253 | 0.9250156 | 0.05335266 |
| **CFA Model (three factor)** |  |  |  |  |  |  |
| F1 =~ NA*is + ihd + t2d + F2 =~ NA*t1d + F3 =~ NA*ldl | 13.40298 | 4 | 0.009465746 | 35.40298 | 0.9343344 | 0.04331982 |
| F1 =~ NA*is + ihd + t2d + bmi + F2 =~ NA*t1d + F3 =~ NA*ldl | 28.30103 | 8 | 0.0004202681 | 54.30103 | 0.9151155 | 0.05563763 |
| F1 =~ NA*is + ihd + t2d + F2 =~ NA*t1d + F3 =~ NA*ldl + bmi | 30.8344 | 7 | 6.67E-05 | 58.8344 | 0.9003415 | 0.05550823 |
| F1 =~ NA*is + t2d + F2 =~ NA*t1d + ihd + F3 =~ NA*ldl + bmi | 22.97458 | 6 | 0.0008050369 | 52.97458 | 0.9290244 | 0.04660928 |
| **Model fit indices for models tested in East Asian population** | | | | | | |
| **CFA Model (one factor)** | **ChiSq** | **df** | **p-value** | **AIC** | **CFI** | **SRMR** |
| F1 =~ NA*is + ihd + hyp + bmi + t2d | 17.33449 | 5 | 0.003907225 | 37.33449 | 0.9287463 | 0.04878802 |
| F1 =~ NA*is + hyp + bmi + t2d | 14.65146 | 2 | 0.0006583793 | 30.65146 | 0.9239025 | 0.05528521 |
| **F1 =~ NA*is + ihd + bmi + t2d** | **1.493882** | 2 | **0.4738138** | 17.49388 | **1** | **0.02240414** |
| **F1 =~ NA*ihd + hyp + bmi + t2d** | **1.35271** | 2 | **0.508467** | 17.35271 | **1** | **0.01226065** |
| **F1 =~ NA*is + ihd + hyp + t2d** | **0.684349** | 2 | **0.7102243** | 16.68435 | **1** | **0.01205982** |
| **F1 =~ NA*is + ihd + hyp + bmi** | **1.958075** | 2 | **0.3756726** | 17.95807 | **1** | **0.01975482** |
| **CFA Model (two factor)** |  |  |  |  |  |  |
| F1 =~ NA*is + hyp + F2 =~ NA*bmi + t2d | **0.2463145** | 1 | **0.6196822** | 18.24631 | **1** | **0.005397** |
| F1 =~ NA*is + hyp + F2 =~ NA*bmi + t2d + F1 | **0.2463145** | 1 | **0.6196822** | 18.24631 | **1** | **0.005397** |
| F1 =~ NA*is + hyp + ihd + F2 =~ NA*bmi + t2d | **2.29661** | 4 | **0.6813863** | 24.29661 | **1** | **0.01744679** |
| F1 =~ NA*is + hyp + ihd + F2 =~ NA*bmi + t2d + F1 | **2.29661** | 4 | **0.6813864** | 24.29661 | **1** | **0.01744679** |
| F1 =~ NA*is + hyp + t2d + F2 =~ NA*bmi + ihd | model failed to converge | | | | | |
| F1 =~ NA*is + hyp + bmi + F2 =~ NA*t2d + ihd | 17.54793 | 4 | 0.001512123 | 39.54793 | 0.9217365 | 0.04841501 |
| F1 =~ NA*is + hyp + ihd + F2 =~ NA*t2d + bmi + ihd | **0.8723346** | 3 | **0.8320985** | 24.87233 | **1** | **0.01018799** |
| F1 =~ NA*is + ihd + F2 =~ NA*t2d + bmi + hyp | model failed to converge | | | | | |
| F1 =~ NA*is + t2d + bmi + F2 =~ NA*ihd + hyp | model failed to converge | | | | | |
| F1 =~ NA*is + t2d + F2 =~ NA*bmi + hyp | model failed to converge | | | | | |
| F1 =~ NA*is + bmi + F2 =~ NA*t2d + hyp | model failed to converge | | | | | |
| **CFA Model (three factor)** | **ChiSq** | **df** | **p-value** | **AIC** | **CFI** | **SRMR** |
| F1 =~ NA*is + hyp + F2 =~ NA*t2d + bmi + F3 =~ NA*ihd | **0.8723344** | 3 | **0.8320985** | 24.87233 | **1** | **0.01018799** |
| F1 =~ NA*is + ihd + F2 =~ NA*t2d + bmi + F3 =~ NA*hyp | model failed to converge | | | | | |
| F1 =~ NA*is + t2d + F2 =~ NA*hyp + bmi + F3 =~ NA*ihd | model failed to converge | | | | | |
| F1 =~ NA*is + bmi + F2 =~ NA*hyp + ihd + F3 =~ NA*t2d | model failed to converge | | | | | |

**Supplementary Table S4. Overlapping genomic risk loci for latent factor F2 between Europe and East Asia**. The table shows the overlapping risk regions between Europe and East Asia for latent factor F2.

| **European region** | **# of significant SNPs** | **# eQTL genes** | **East Asian region** | **# of significant SNPs** | **# eQTL genes** |
| --- | --- | --- | --- | --- | --- |
| 1:177,803,163-177,913,519 | 1 | 0 | 1:177,843,479-177,936,599 | 9 | 0 |
| 4:45,165,650-45,187,622 | 1 | 0 | 4:45,163,333-45,187,622 | 3 | 0 |
| 6:50,787,595-50,940,677 | 1 | 0 | 6:50,644,216-50,940677 | 5 | 0 |
| 11:27,541,623-27,242,447 | 1 | 1 | 11:27,634,373-27,749,725 | 5 | 2 |
| 12:50,179,021-50,275,385 | 1 | 1 | 12:52,261,809-50,285,780 | 2 | 1 |
| 16:53,755,146-53,848,561 | 3 | 1 | 16:53,765,639-53,848,561 | 15 | 0 |
| 18:57,728,947-58,017,249 | 7 | 0 | 18:57,715,582-58,009,426 | 11 | 0 |

(A) European dataset	(B) East Asian dataset

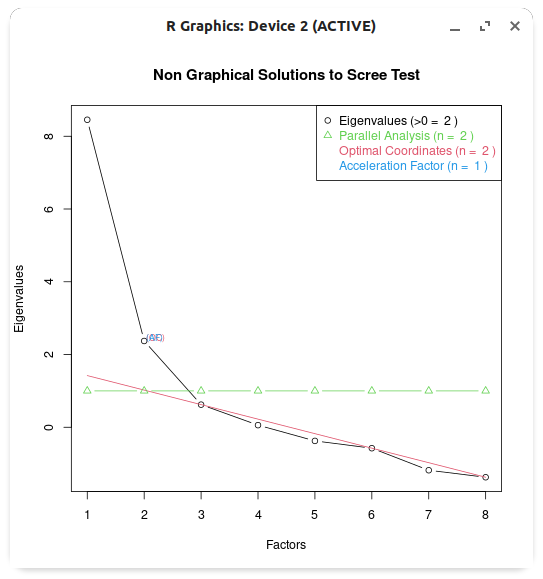

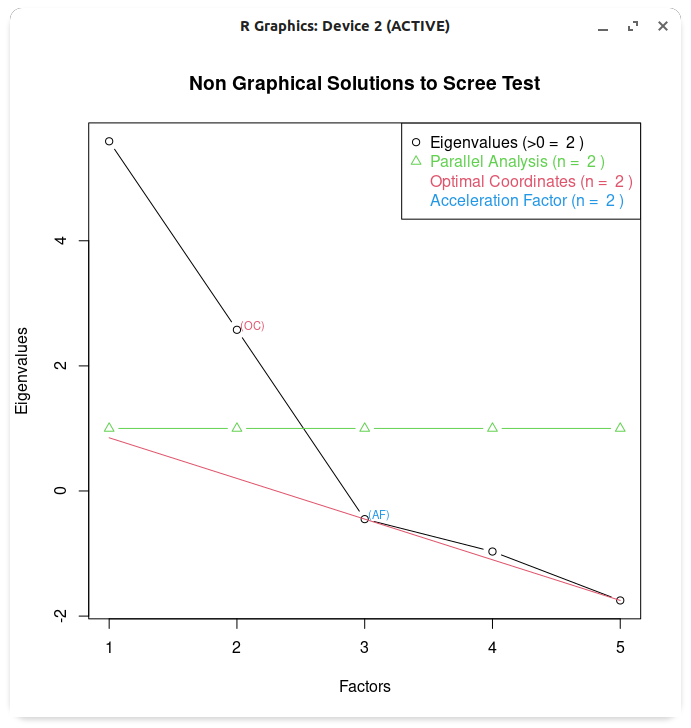

**Supplementary Figure S1. Scree plot from exploratory factor analysis indicating optimal number of factors for (A) European and (B) East Asian summary statistics.** The different methods indicate that both the European and East Asian datasets optimally have two factors each.

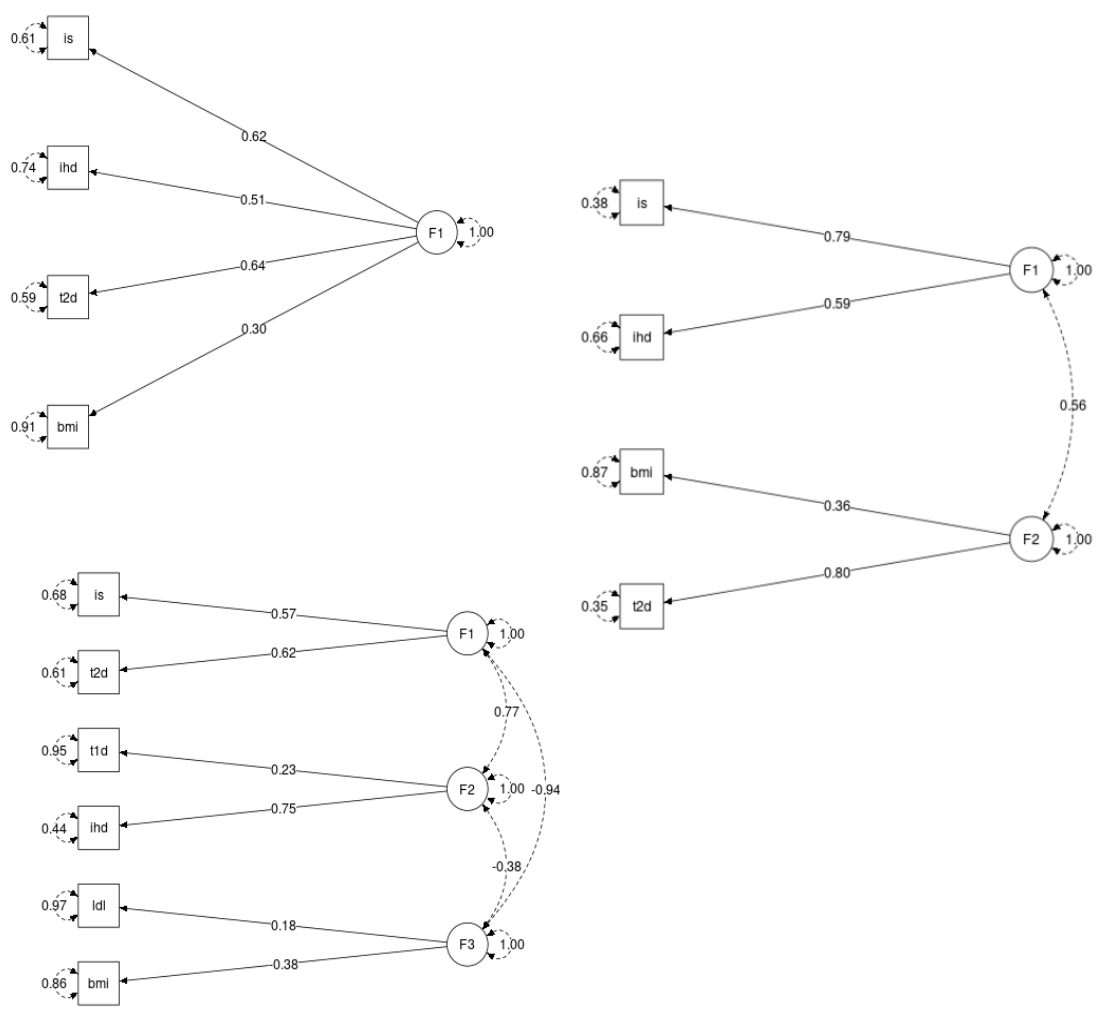

**Supplementary Figure S2A. Confirmatory factor analysis models of stroke and its comorbid conditions with one factor, two factor and three factors in the European population.** The models tested in confirmatory factor analysis with one, two and three latent factors among stroke and its comorbid conditions with factors loadings and correlations between them.

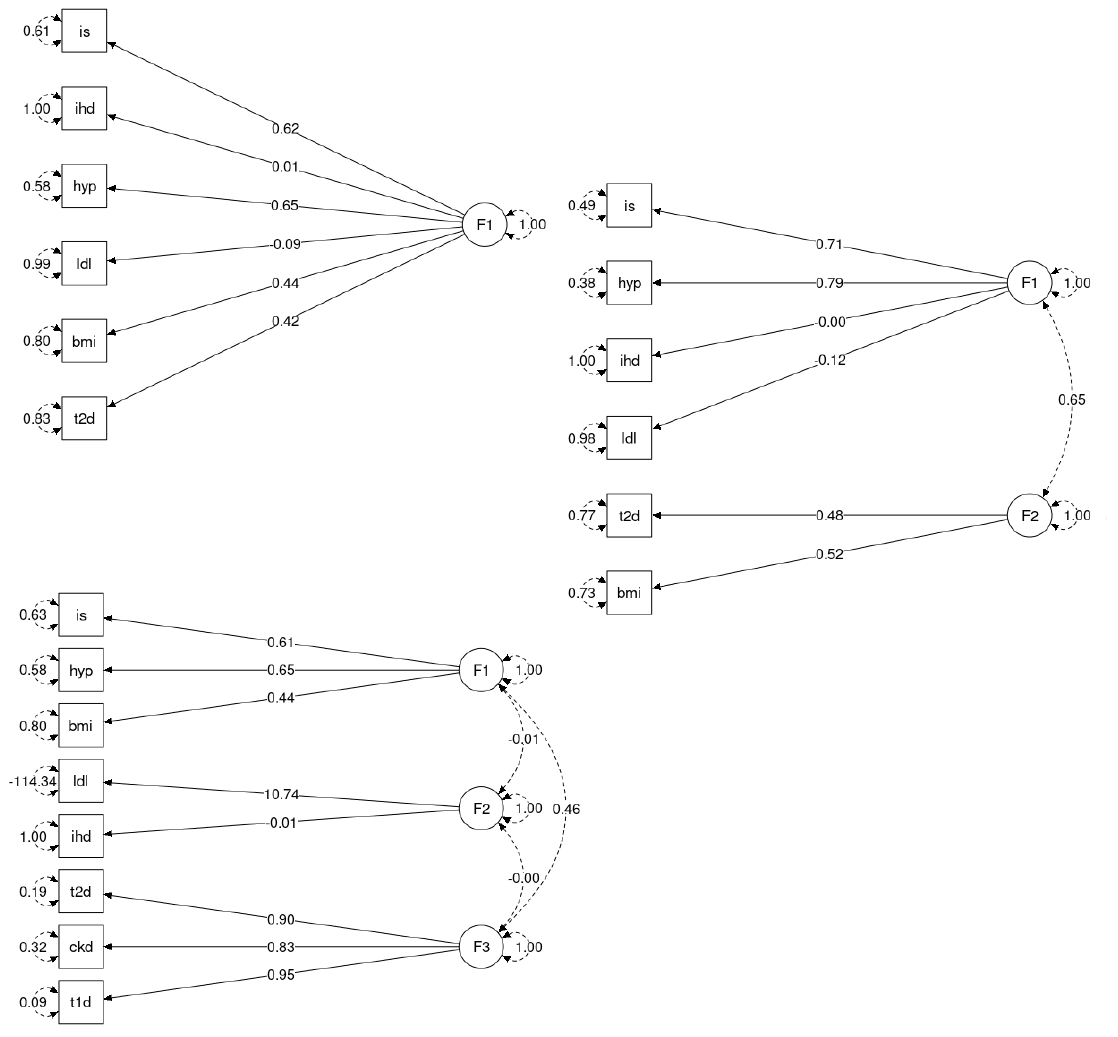

**Supplementary Figure S2B. Confirmatory factor analysis models of stroke and its comorbid conditions with one factor, two factor and three factors in the East Asian population.** The models tested in confirmatory factor analysis with one, two and three latent factors among stroke and its comorbid conditions with factors loadings and correlations between them.

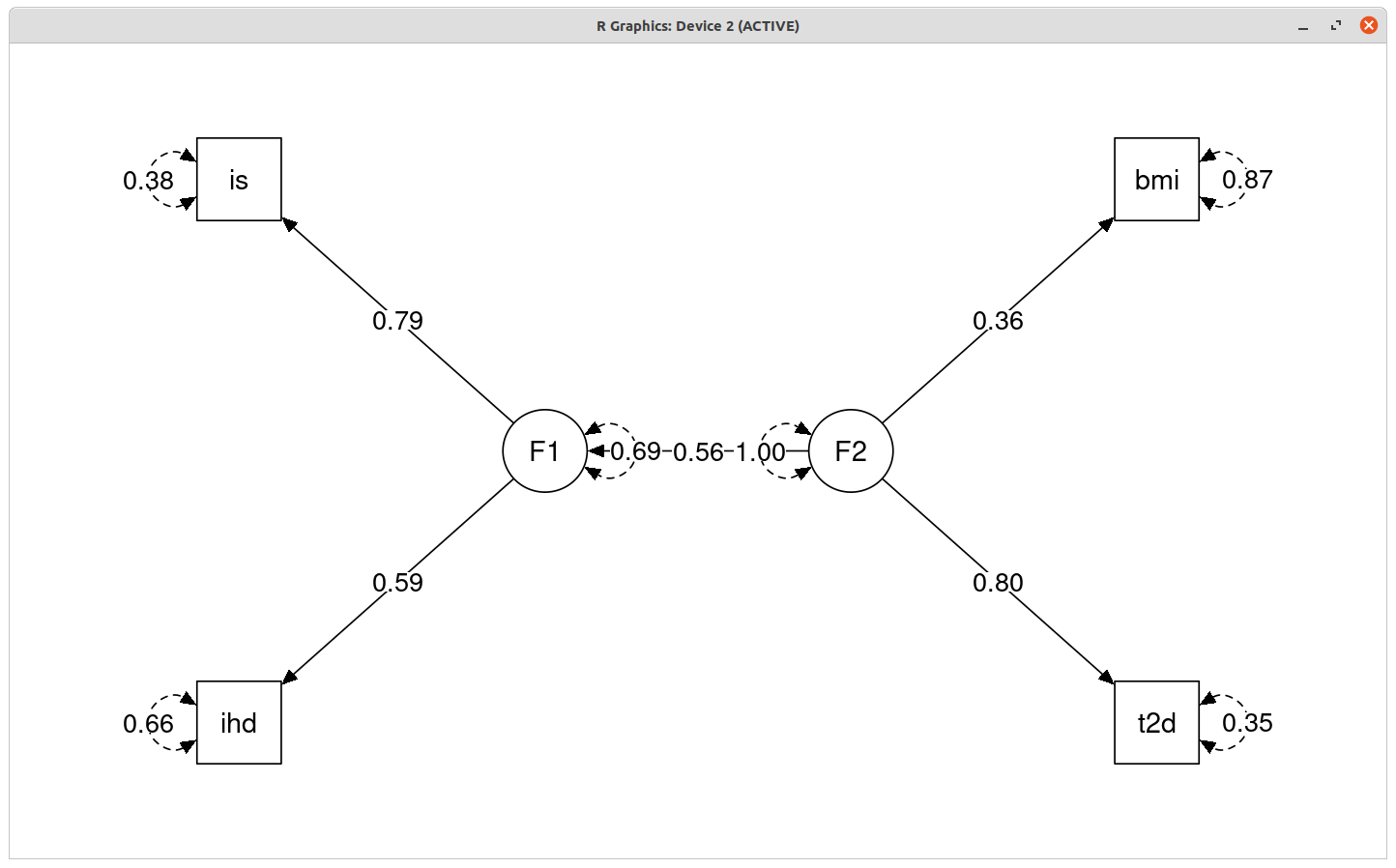

**Supplementary Figure S3. Secondary genetic models proposed for stroke and its comorbid conditions in the European population.** The second proposed genetic models for stroke and its comorbid conditions have two latent factors. Ischemic stroke (is) and ischemic heart disease (ihd) loads onto the first factor F1, and high body mass index (bmi), type 2 diabetes (t2d) as well as F1 loads onto the second factor F2. Both factors, F1 and F2, have path coefficients to each variable greater than 0.3. Factor 1 also loads on factor 2 with a path coefficient of 0.56. The residuals of the variables are shown in circular self-paths.

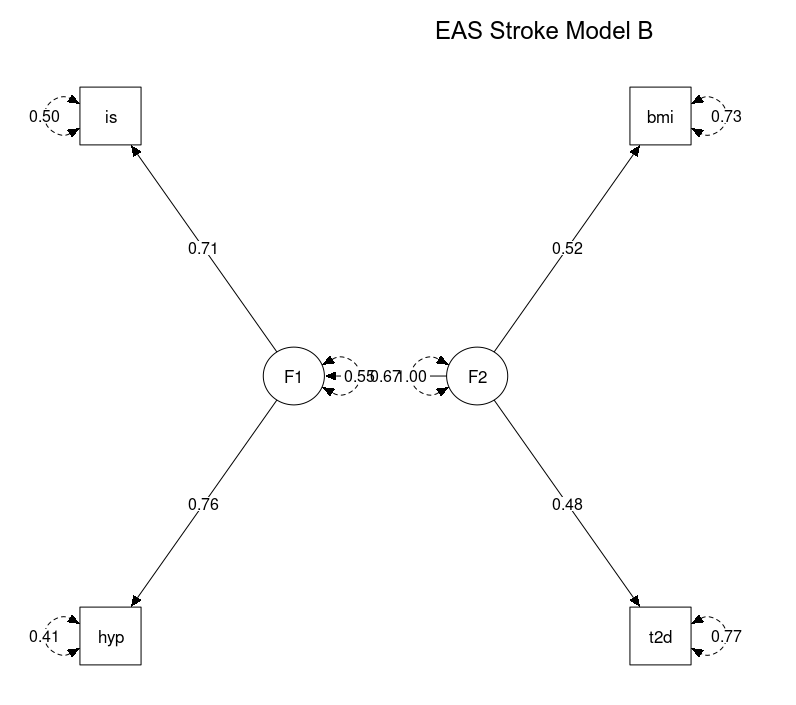

**Supplementary Figure S4. Secondary genetic models proposed for stroke and its comorbid conditions in the East Asian population.** The second proposed genetic models for stroke and its comorbid conditions have two latent factors. Ischemic stroke (is) and high systolic blood pressure (hyp) loads onto the first factor F1, and high body mass index (bmi), type 2 diabetes (t2d) as well as F1 loads onto the second factor F2. Both factors, F1 and F2, have path coefficients to each variable greater than 0.3. Factor 1 also loads on factor 2 with a path coefficient of 0.67. The residuals of the variables are shown in circular self-paths.

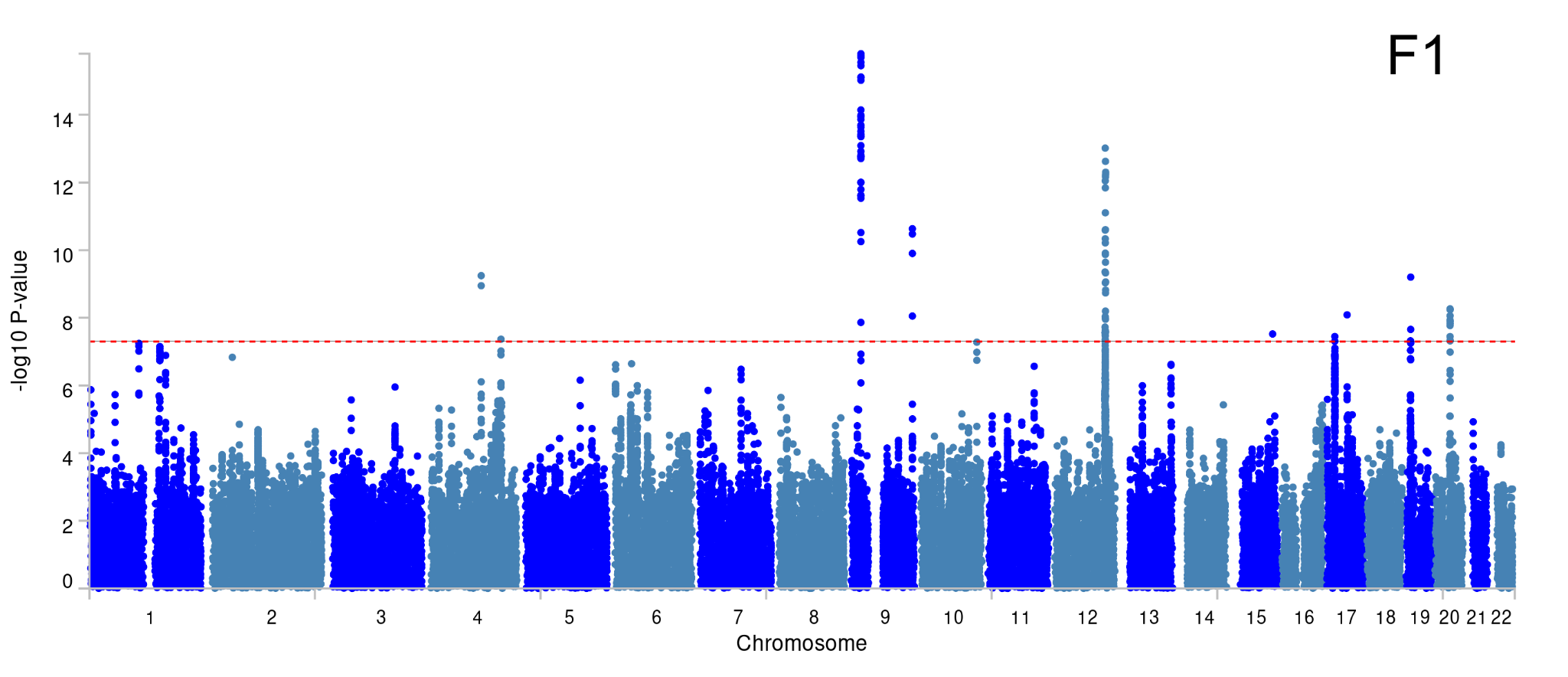

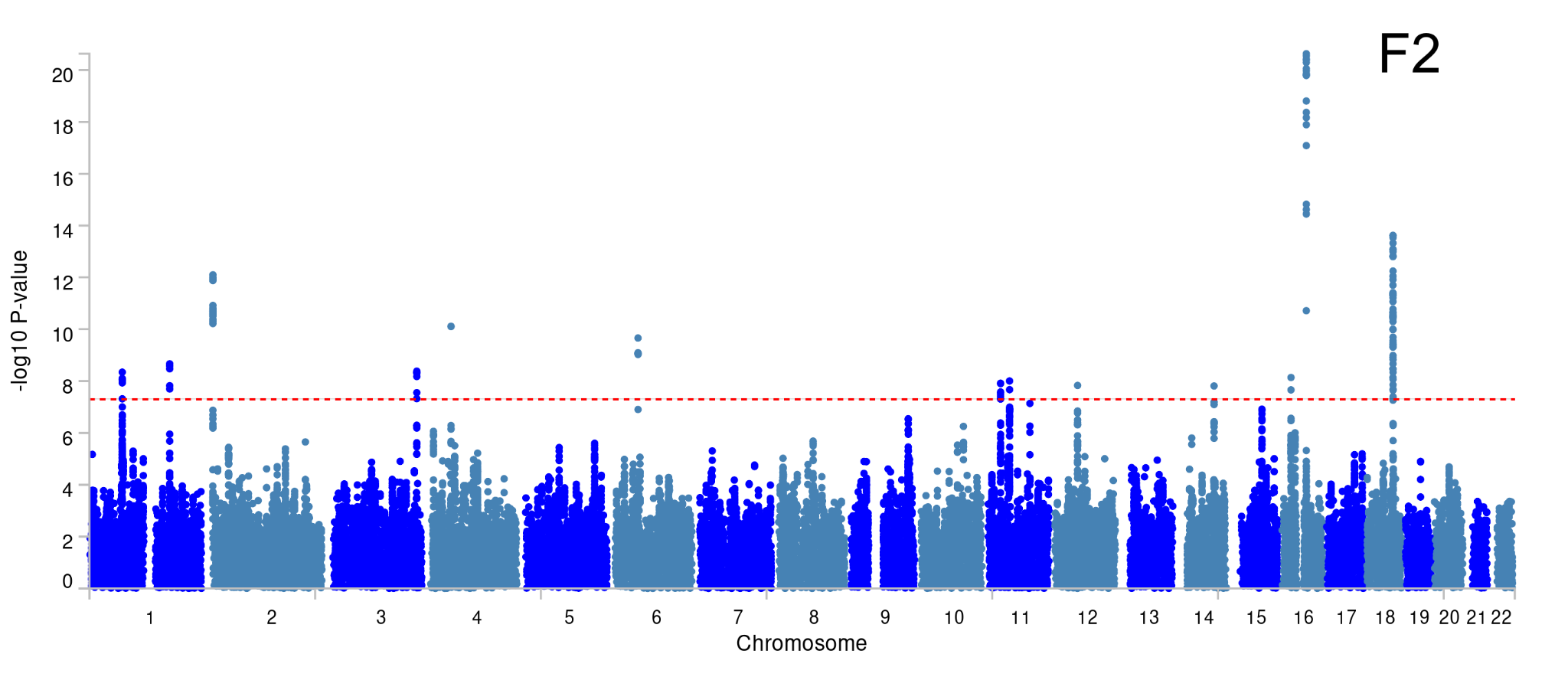

**Supplementary Figure S5. Manhattan plot of GWAS of two latent factors F1 and F2 of stroke and its comorbid conditions in European population.** The plot shows all the SNPs analysed for each factor F1 and F2 as a dot. The X-axis shows the chromosome location of the SNP, the y-axis shows the negative log of p-value. SNPs above the threshold line of 5e-08 are considered significant.

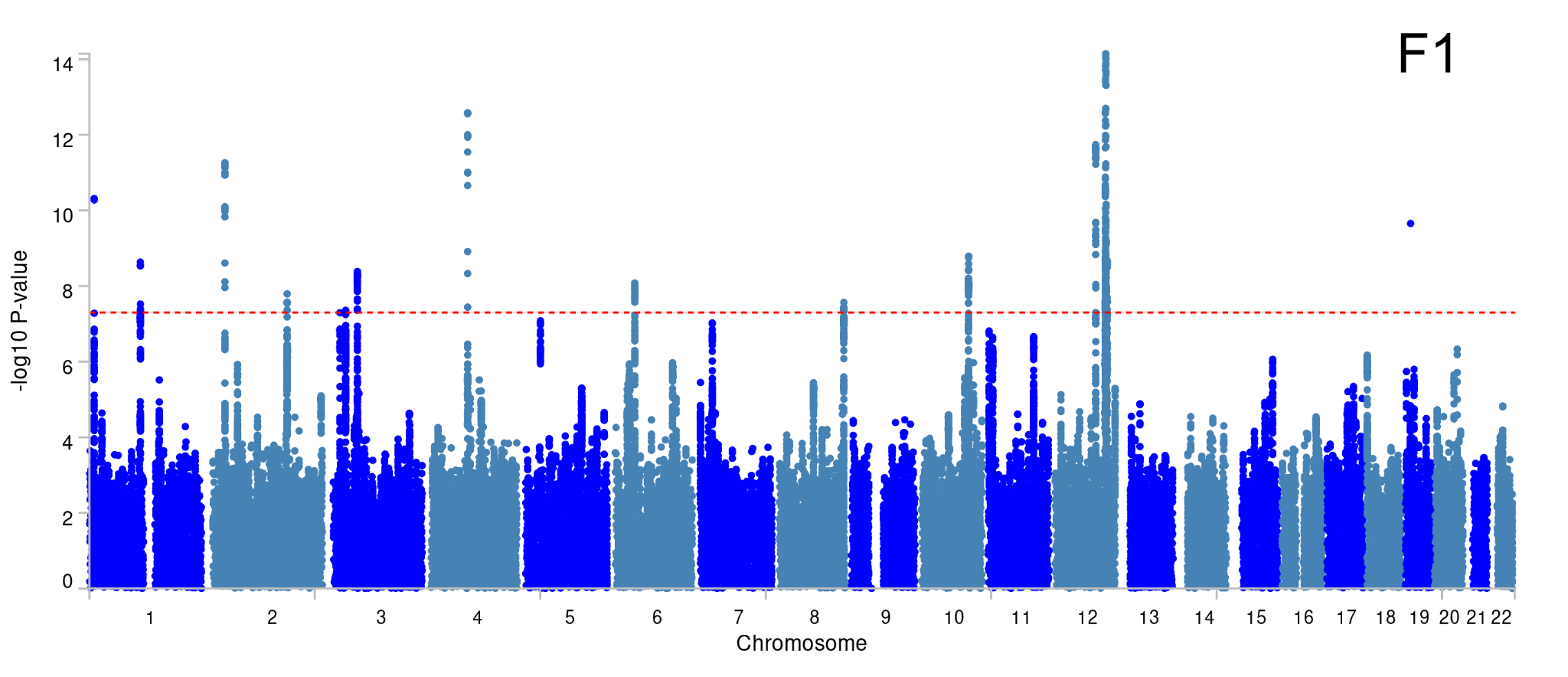

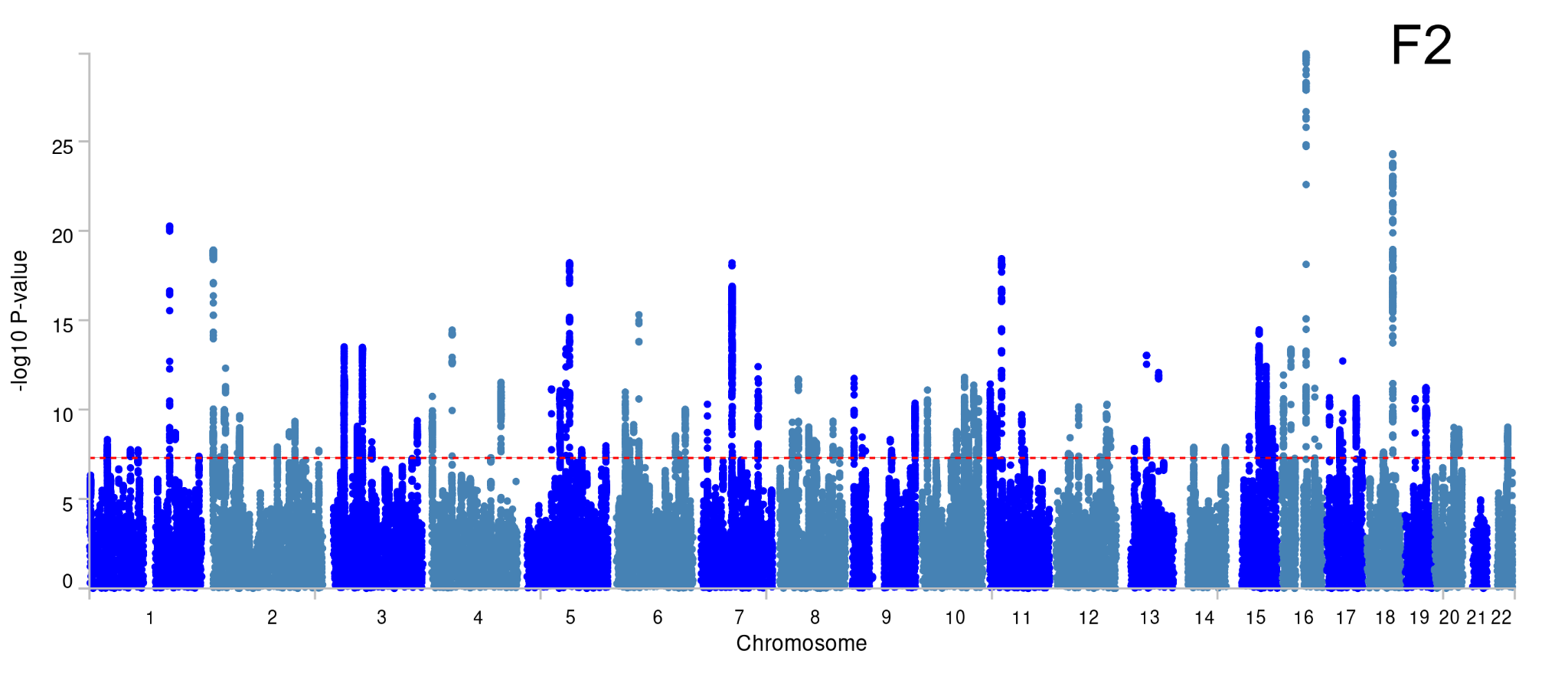

**Supplementary Figure S6. Manhattan plot of GWAS of two latent factors F1 and F2 of stroke and its comorbid conditions in East Asian population.** The plot shows all the SNPs analysed for each factor F1 and F2 as a dot. The X-axis shows the chromosome location of the SNP, the y-axis shows the negative log of p-value. SNPs above the threshold line of 5e-08 are considered significant.

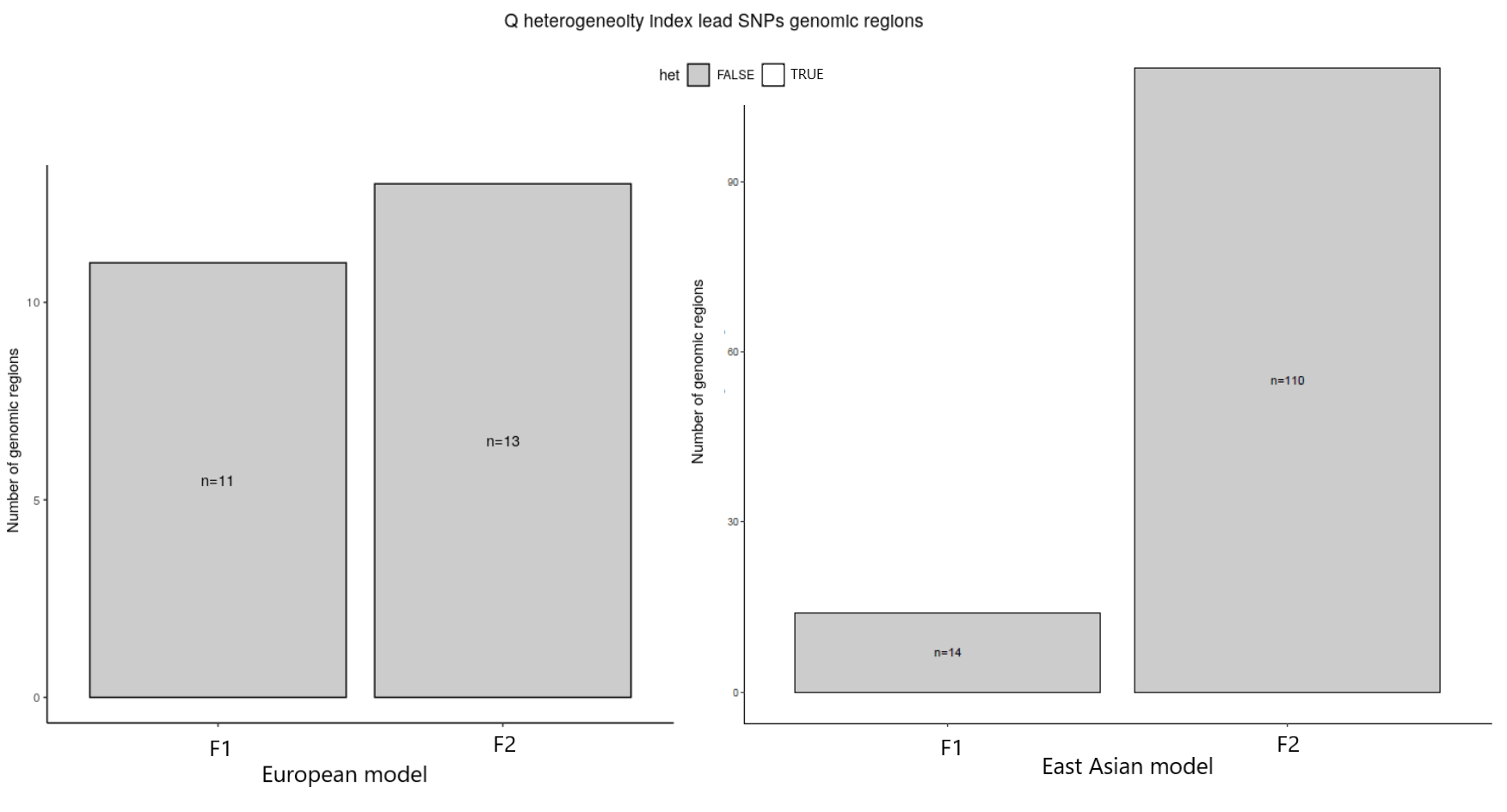

**Supplementary Figure S7. Q_SNP_ heterogeneity test per genomic risk locus of latent factors F1 and F2 in the European and East Asian models.** The chi-square tests the null hypothesis that the SNPs in the genomic risk loci act through the defined model. Hence, if significant, the SNPs are indeed heterogeneous and the test returns TRUE. If non-significant, the SNPs are acting only through our model, and thus, the test returns FALSE.

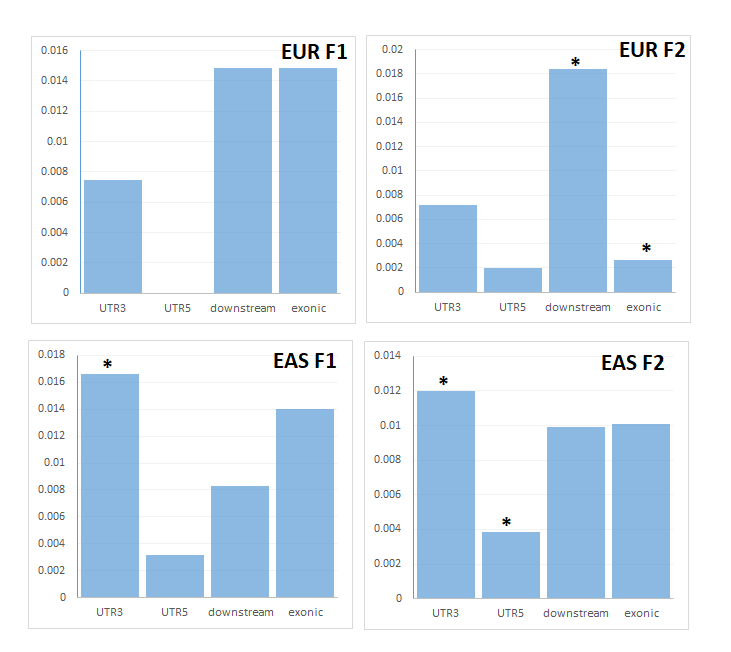

**Supplementary Figure S8. The gene location (exonic, downstream and UTR regions) of the SNPs associated with the latent factors.** The histogram displays the proportion of all SNPs in LD with significant SNPs which have corresponding functional annotation from ANNOVAR. Bars are coloured by log2(enrichment) relative to all SNPs selected in the reference panel. EUR F1 and EUR F2 are factors in European population and EAS F1 and EAS F2 are factors in East Asian population.
